# Transfer RNA fragments replace microRNA regulators of the cholinergic post-stroke immune blockade

**DOI:** 10.1101/2020.07.02.20144212

**Authors:** Katarzyna Winek, Sebastian Lobentanzer, Bettina Nadorp, Serafima Dubnov, Claudia Dames, Sandra Jagdmann, Gilli Moshitzky, Benjamin Hotter, Christian Meisel, David S Greenberg, Sagiv Shifman, Jochen Klein, Shani Shenhar-Tsarfaty, Andreas Meisel, Hermona Soreq

**Affiliations:** The Edmond & Lily Safra Center for Brain Sciences, The Hebrew University of Jerusalem, Jerusalem, Israel; The Alexander Silberman Institute of Life Sciences, The Hebrew University of Jerusalem, Jerusalem, Israel; Department of Pharmacology, College of Pharmacy, Goethe University, Frankfurt am Main, Germany; The Grass Center for Bioengineering, Benin School of Computer Science and Engineering, The Hebrew University of Jerusalem, Jerusalem, Israel; The Institute for Medical Immunology, Charité-Universitätsmedizin Berlin, Germany; NeuroCure Clinical Research, Center for Stroke Research Berlin and The Department of Neurology, Charité-Universitätsmedizin Berlin, Germany; The Department of Genetics, The Hebrew University of Jerusalem, Jerusalem, Israel; Department of Internal Medicine “C”, “D” and “E”, Tel Aviv Sourasky Medical Center and Sackler Faculty of Medicine, Tel Aviv University, Tel Aviv, Israel

**Keywords:** acetylcholine, ischemic stroke, microRNA, immunology, transfer RNA fragment

## Abstract

Stroke is a leading cause of death and disability. Recovery depends on a delicate balance between inflammatory responses and immune suppression, tipping the scale between brain protection and susceptibility to infection. Peripheral cholinergic blockade of immune reactions fine-tunes this immune response, but its molecular regulators are unknown. Here, we report a regulatory shift in small RNA types in patient blood sequenced two days after ischemic stroke, comprising massive decreases of microRNA levels and concomitant increases of transfer RNA fragments (tRFs) targeting cholinergic transcripts. Electrophoresis-based size-selection followed by RT-qPCR validated the top 6 upregulated tRFs in a separate cohort of stroke patients, and independent datasets of small and long RNA sequencing pinpointed immune cell subsets pivotal to these responses, implicating CD14^+^ monocytes in the cholinergic inflammatory reflex. In-depth small RNA targeting analyses revealed the most-perturbed pathways following stroke and implied a structural dichotomy between microRNA and tRF target sets. Furthermore, lipopolysaccharide stimulation of murine RAW 264.7 cells and human CD14^+^ monocytes upregulated the top 6 stroke-perturbed tRFs, and overexpression of stroke-inducible tRF-22-WE8SPOX52 using an ssRNA mimic induced downregulation of immune regulator Z-DNA binding protein 1 (Zbp1). In summary, we identified a “changing of the guards” between RNA types that may systemically affect homeostasis in post-stroke immune responses, and pinpointed multiple affected pathways, which opens new venues for establishing therapeutics and biomarkers at the protein- and RNA-level.

**Significance Statement:** Ischemic stroke triggers peripheral immunosuppression, increasing the susceptibility to post-stroke pneumonia that is linked with poor survival. The post-stroke brain initiates intensive communication with the immune system, and acetylcholine contributes to these messages; but the responsible molecules are yet unknown. We discovered a “changing of the guards,” where microRNA levels decreased but small transfer RNA fragments (tRFs) increased in post-stroke blood. This molecular switch may re-balance acetylcholine signaling in CD14^+^ monocytes by regulating their gene expression and modulating post-stroke immunity. Our observations point out to tRFs as molecular regulators of post-stroke immune responses that may be potential therapeutic targets.

## Main Text

### Introduction

Stroke is a global burden of growing dimensions, accounting for ca. 5.5 million deaths annually, and leaving most of the surviving patients permanently disabled (1). The immune system is one of the main players in the pathophysiology of stroke. Brain injury dampens immune functions in the periphery, which limits the inflammatory response and infiltration of immune cells into the CNS and may pose a neuroprotective mechanism in stroke patients. However, this systemic immunosuppression simultaneously increases the risk of infectious complications (2), e.g. by inducing lymphocyte apoptosis and decreasing the production of pro-inflammatory cytokines (lymphocytic IFNγ and monocytic TNFα) (3). Therefore, post-stroke recovery largely depends on a delicate balance between inflammation, which exacerbates the severity of symptoms, and the post-stroke suppression of immune functions, which increases the susceptibility to infections (3). This involves incompletely understood molecular regulator(s) of cholinergic and sympathetic signaling and the hypothalamus-pituitary-adrenal gland axis (HPA). Among other processes, brain injury leads to activation of the vagus nerve, which mediates anti-inflammatory signaling through the cholinergic efferent fibers and the noradrenergic splenic nerve (4). Binding of acetylcholine (ACh) to the nicotinic alpha 7 receptors on monocytes/macrophages decreases the production of proinflammatory cytokines (4, 5) in a manner susceptible to suppression by microRNA (miR) regulators of cholinergic signaling, such as miR-132 (6). We hypothesized that this and other small RNA fine-tuners of innate immune responses, including miRs and the recently re-discovered transfer RNA (tRNA) fragments (tRFs), may contribute to regulation of post-stroke processes.

Both miRs and tRFs may control entire biological pathways, such that their balanced orchestration could modulate brain-induced systemic immune functioning. miRs are small non-coding RNAs whose expression requires transcription yet can be rapidly induced, enabling degradation and/or translational suppression of target genes carrying a complementary motif. One miR may suppress the expression of many targets involved in the same biological pathway, and many miRs may co-target the same transcripts, enabling cooperative suppression. Hence, miR regulators of ACh signaling may regulate the role of ACh in both cognitive function and systemic inflammation (6, 7).

Recent reports highlight tRNA as another major source of small noncoding RNA(8), including tRNA halves (tiRNAs) and smaller tRNA fragments - tRFs. tiRNAs are created by angiogenin cleavage at the anticodon loop (9) raising the possibility that the post-stroke angiogenin increase might change their levels (10). Among other functions (11, 12), smaller fragments derived from the 3’-or 5’-end of tRNA (3’-tRF/5’-tRF) or internal tRNA parts (i-tRF) may incorporate into Argonaute (Ago) protein complexes and act like miRs to suppress their targets (13). Differential expression of tRFs was reported under hypoxia, oxidative stress, ischemic reperfusion (9, 14) and in epilepsy (15), which are all involved in ischemic stroke complications. tRFs may be generated via enzymatic degradation of tRNA, independent of de-novo transcription, which implies that tRF levels may be modulated more rapidly than miR levels. However, whether brain-body communication and immune suppression after ischemic stroke in human patients involves blood tRF changes has not yet been studied.

Taking into consideration that the cholinergic system is one of the controllers of immune functions, we investigated changes in the levels of miR- and tRF-regulators, with a specific focus on those which may control the ACh-mediated suppression of post-stroke immune functions. We performed small and long RNA-sequencing of whole blood samples collected from ischemic stroke patients two days after stroke onset, mined RNA-sequencing datasets of blood cell transcripts and sought potential links between perturbed miRs and tRFs, post-stroke immune responses and the cholinergic anti-inflammatory pathway.

### Results

#### Stroke-perturbed small RNAs display a cholinergic-associated shift from miRs to tRFs

To seek post-stroke small RNA regulators of body-brain communication, we first performed small RNA-sequencing of whole blood samples collected on day 2 after ischemic stroke from 33 male patients of the PREDICT cohort (484 participants) (16) and 10 age- and sex-matched controls (Fig. 1A; see demographic data in Data file S1). Principal Component Analysis (PCA) of the differentially expressed (DE) small RNAs completely segregated the stroke and control groups (Fig. 1B). The respective direction of change among the two small RNA classes involved a statistically significant decline in miRs and a parallel increase in tRFs, indicating a ‘changing of the guards’ from miRs to tRFs. Specifically, 87% of the 143 DE tRFs were upregulated, whereas 63% of the 420 DE miRs were downregulated (Benjamini-Hochberg corrected p < 0.05; Fig. 1C&D). Of the 143 DE tRFs, 87 were 3’-tRFs, and 30 of those (all upregulated) were derived from alanine binding tRNA (Fig. S1), indicating non-arbitrary fragment generation.

**Figure 1.**
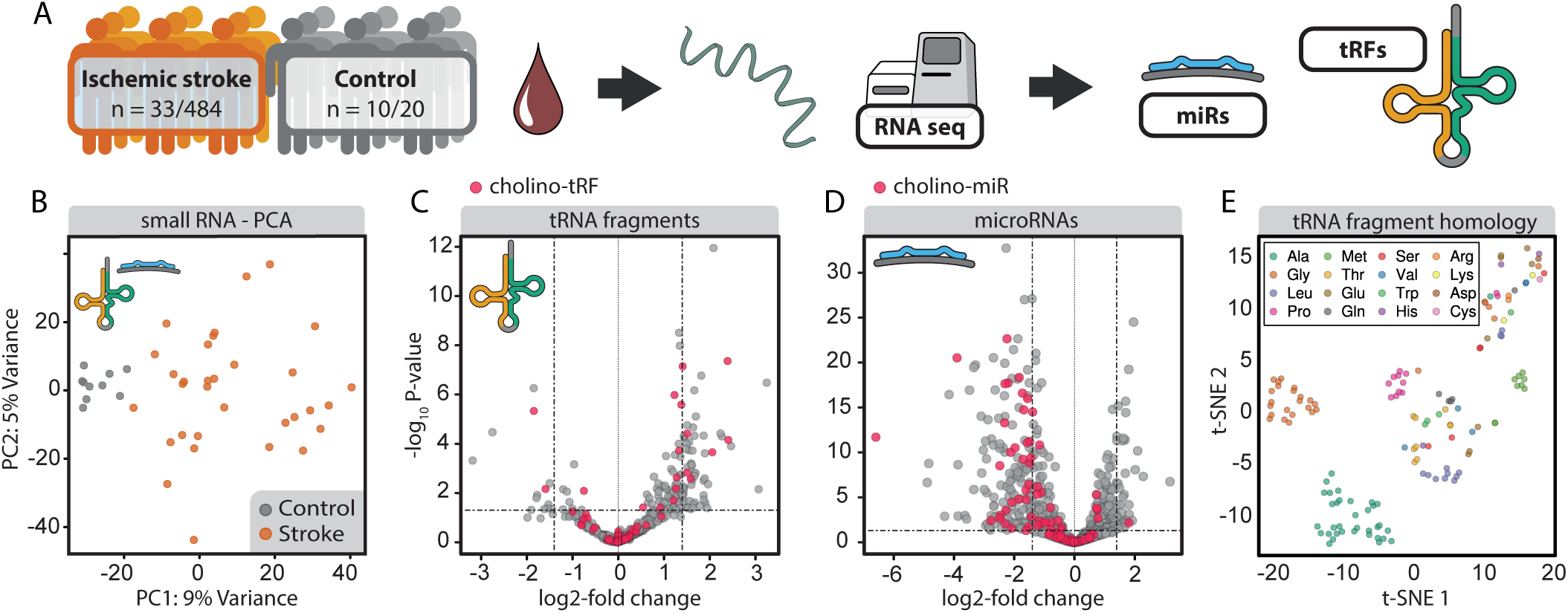
Post-stroke differential expression of small RNA species and tRF homology clustering. A) Whole blood total RNA were collected on day 2 post-stroke from patients of the PREDICT cohort (NCT01079728) (16) and age-matched controls. B) PCA of DE tRFs/miRs in patients’ blood separated stroke and control samples. C) Volcano plot of DE tRFs from stroke patients and controls (horizontal line at adjusted p = 0.05) showing upregulation of most DE tRFs. D) Volcano plot of DE miRs shows predominant downregulation in stroke patients compared with controls (horizontal line at adjusted p = 0.05). Red dots in C and D reflect Cholino-tRFs and Cholino-miRs, respectively. E) t-SNE visualization of tRF homology based on pairwise alignment scores of sequences of all detected tRNA fragments shows grouped tRFs of several specific amino acid origins.

Notably, the 420 DE miRs included several miRs known to be perturbed in stroke: hsa-miR-532-5p (logFC = −2.27, p = 1.81e-33) (17), hsa-miR 148a-3p (logFC = −2.30, p = 9.61e-19) and hsa-let-7i-3p (logFC = −1.07, p = 4.31e-04) (18). To test the potential involvement of miRs and tRFs in regulating the cholinergic anti-inflammatory pathway after stroke, we performed targeting analysis of DE miRs and tRFs towards cholinergic transcripts (Supplementary Methods, for a complete list of cholinergic genes see Data File S2) via an in-house integrative database (*miRNeo*) (19) containing comprehensive transcription factor (TF)- and miR-targeting data, complemented by de-novo prediction of tRF-targeting using TargetScan (20). A restrictive approach identified 131 miRs and 64 tRFs containing complementary motifs to at least five cholinergic-associated transcripts each (further termed “Cholino-miRs” and “Cholino-tRFs”, Fig. 1C&D, Fig. S2, full lists in Data files S3 and S4). Permutation targeting analysis showed an enrichment of cholinergic targets for both DE miRs (100 000 permutations, p = 0.0036) and DE tRFs (100 000 permutations, p = 2e-05). Further indicating non-random generation of these fragments, the tRFs identified in our dataset clustered into oligonucleotide families with high sequence homology via t-distributed stochastic neighbor embedding (t-SNE) (Fig. 1E), including families known to associate with Ago and suppress growth and proliferation via post-transcriptional downregulation in lymphocytes (e.g. tRF-22-WE8SPOX52 from tRNAGly) (21) and metastatic cancer cells (tRF-18-HR0VX6D2 from tRNALeu) (22). This supported our prediction that the concomitant elevation of tRF- and decline of miR-levels in post-stroke blood could contribute to the post-stroke changes in cholinergic signaling pathways.

To further challenge our findings, we validated the expression levels of prominently DE tRFs identified by RNA sequencing in a separate cohort of PREDICT patients (16). Standard qPCR techniques cannot distinguish between the full length tRNA molecules and their 3’-tRF cleavage products. Therefore, to experimentally validate tRF changes (Fig. 2A) we implemented an electrophoresis size selection-based strategy followed by cDNA synthesis from the selected small RNAs and RT-qPCR (maximum 25 nt, Fig. 2B). This procedure validated the top six upregulated tRFs identified in RNA-sequencing data (tRF-22-WEKSPM852, tRF-18-8R6546D2, tRF-18-HR0VX6D2, tRF-18-8R6Q46D2, tRF-22-8EKSP1852 and tRF-22-WE8SPOX52; according to count-change, see Supplementary Methods, Fig. 2B&C) and demonstrated significant increases accompanied by higher variability in the blood levels of these tRFs in post-stroke patients compared to controls.

**Figure 2.**
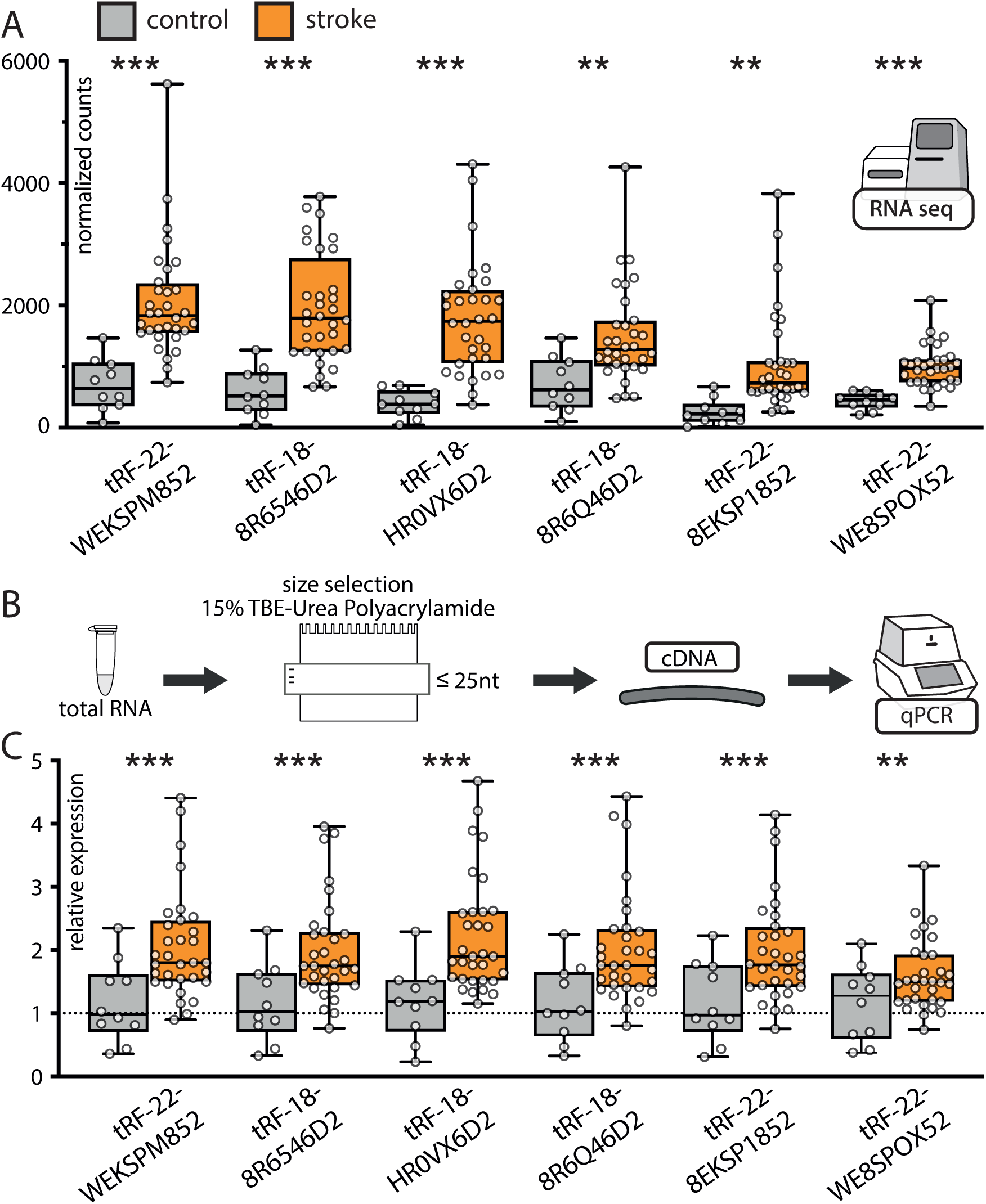
RT-qPCR validation of the top 6 upregulated tRFs in PREDICT stroke patients following size selection for small RNA. A) RNA-sequencing counts normalized to the size of the library (using DESeq2 (23)) of the top 6 upregulated tRFs (from left to right). Asterisks indicate adjusted p-values of Wald test via DESeq2, ** p < 0.01, *** p < 0.001, shown are box-plots with whiskers minimum to maximum. B) Size selection workflow for validations in a separate sub-group of PREDICT stroke patients (n=32) using the same control group (n=10); C) RT-qPCR validations using normalized expression (hsa-miR-30d-5p, hsa-let7d-5p, hsa-miR-106b-3p and hsa-miR-3615 served as house-keeping transcripts, see Supplementary Methods), relative to the control group (line at mean normalized expression for the control group =1) confirmed upregulation of top 6 DE tRFs identified in RNA-sequencing, one-way ANOVA, ** p < 0.01, *** p < 0.001, box-plots with whiskers minimum to maximum.

#### Stroke-perturbed whole blood tRFs are biased towards cellular blood compartments

To clarify the distribution of stroke-perturbed tRFs among the immunologically relevant blood cell types, we mined an RNA-sequencing dataset comprising sorted cell populations collected from healthy volunteers: CD4^+^ T helper cells, CD8^+^ T cytotoxic cells, CD56^+^ NK cells, CD19^+^ B cells, CD14^+^ monocytes, CD15^+^ neutrophils, CD235a^+^ erythrocytes, serum, exosomes, and whole blood (450 samples in total, (24)) (Fig. 3A). Predicting that log-normal distribution of the counts in different samples would point towards biological significance, we categorized all tRFs found in this dataset into present/absent in a specific blood compartment (without introducing a limit for counts, see Methods, Fig. 3B). Two main clusters of specific blood compartments could be identified based on their specific tRF profile: a) monocytes, B-, T-, and NK cells; b) neutrophils, whole blood, serum, exosomes, and erythrocytes (Fig. S3). Further, we distinguished eight tRF sub-clusters, based on the presence/absence of specific tRFs in blood compartments (Fig. 3C), with cluster four comprising molecules expressed specifically in monocytes, B-, T-, and NK cells, and cluster seven consisting of tRFs expressed only in monocytes.

**Figure 3.**
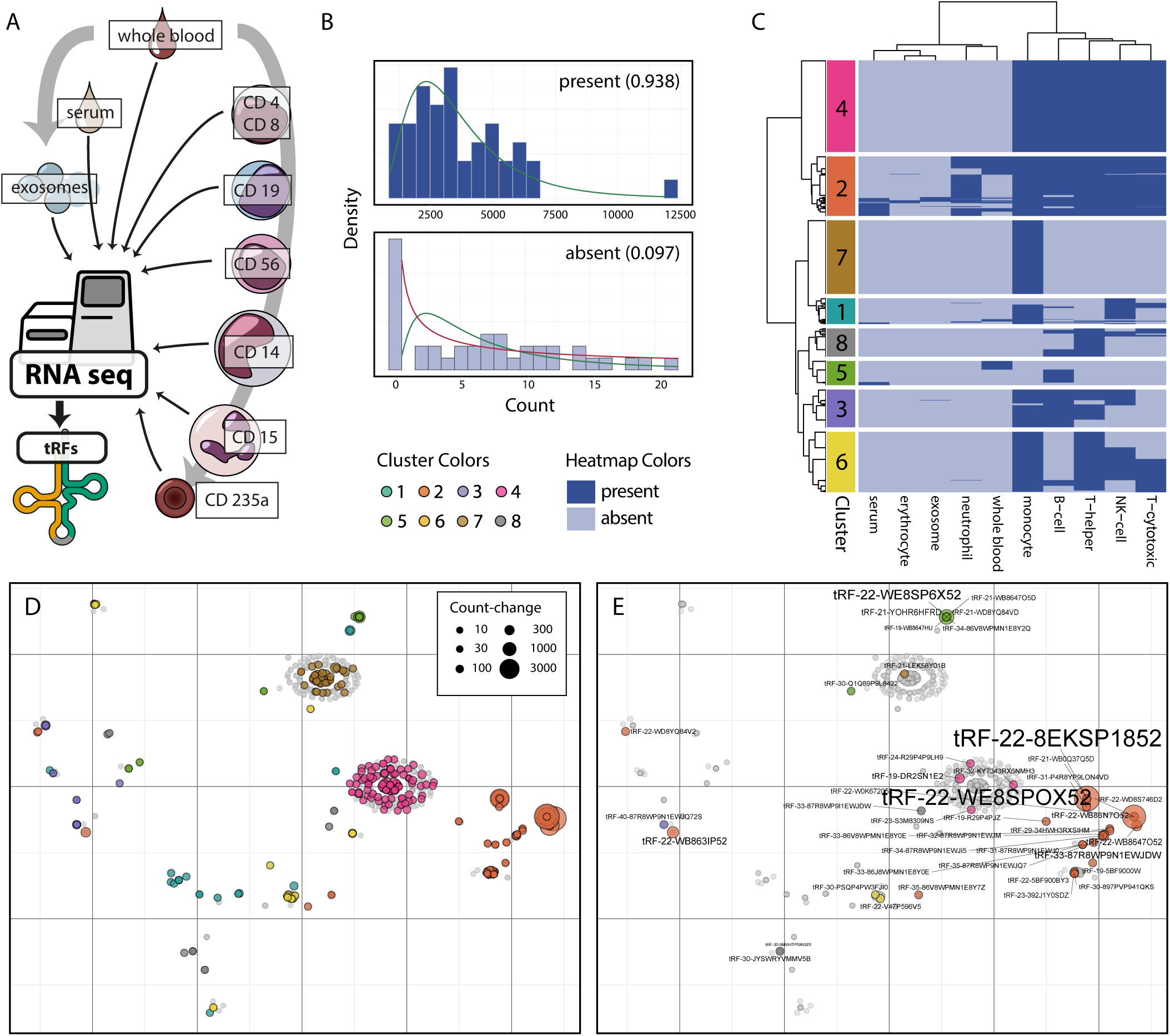
Immune cell tRF expression clustering and cell type-specific analysis. A) Analysis of RNA-sequencing datasets from T lymphocytes (CD4^+^ T helper cells and CD8^+^ T cytotoxic cells), B lymphocytes (CD19^+^), NK cells (CD56^+^), monocytes (CD14^+^), neutrophils (CD15^+^) erythrocytes (CD235a^+^), serum, exosomes, and whole blood (24) yielded a blood tRF profile. B) Definition of presence/absence of small RNAs in these blood compartments via statistical assertion of log-normal count distribution (values between 0 and 1, closer to 1: present). C) Detailed analysis of identified tRFs found 8 sub-clusters based on cell types expressing specific molecules. D) t-SNE of all found tRFs represented by grey dots, DE tRFs identified in the PREDICT study are marked with cluster-specific color. E) t-SNE of all tRFs found, Cholino-tRFs identified in the PREDICT study are marked with cluster-specific color.

Based on this methodology, we conducted a census of small RNA species found intra-vs extracellularly: we detected 1624 distinct intracellular tRFs but only 93 extracellular tRFs; 149 in whole blood, but 1417 in CD14^+^ monocytes alone. Similarly, we detected 559 distinct intracellular miRs but 145 extracellular miRs; 475 in monocytes alone, and 331 in whole blood. Using the presence/absence measure for analyzing the post-stroke DE tRFs (Fig. 3D, Fig. S4 for the top 20 stroke DE tRFs), we detected 77 DE tRFs from the PREDICT dataset as expressed in immune cells (Fig. 3E, a detailed list of tRFs and affiliated clusters in Data File S5), including 10 Cholino-tRFs. Notably, tRFs previously shown to function post-transcriptionally in a miR-like manner (e.g. tRF-22-WE8SPOX52 from tRNAGly (21) and tRF-18-HR0VX6D2 from tRNALeu, alias hsa-miR-1280 (22)) segregated into whole blood, monocyte, T, B- and NK cell compartments rather than into erythrocyte, serum or exosome compartments. Thus, the post-stroke modified tRFs may be functionally involved in regulating the leukocytic post-stroke response.

#### CD14^+^ monocytes show highest cholinergic-related transcriptional repertoire

The enrichment of DE Cholino-miRs and Cholino-tRFs identified in the PREDICT dataset and the contribution of the cholinergic anti-inflammatory pathway to peripheral immunosuppression called for pinpointing the immune compartment(s) in which these small RNAs might affect post-stroke immune suppression. Analysis of long RNA regulatory circuits (25) specific to blood-borne leukocytes (Fig. 4A) identified CD14^+^ monocytes as the main cell type expressing cholinergic core and receptor genes (Data File S2, Fig. 4B). To confirm the relevance of this effect, we performed long RNA-sequencing in blood samples from 20 stroke patients from the PREDICT study and 4 controls. This showed 204 upregulated and 490 downregulated long RNA transcripts. Gene ontology (GO) enrichment analyses of the most implicated genes yielded highly specific terms relevant to innate immunity, vascular processes, and cholinergic links (Fig. 4C, list of all significant terms in Table S1). More specifically, terms linked to innate immune processes in post-stroke blood involved responses to LPS mediated by interferons and other cytokines (Fig. 4C, left-hand side); vascular processes comprised platelet activation and degranulation, control of cell-cell adhesion, and regulation of angiogenesis (Fig. 4C, right-hand side). Intriguingly, differentially regulated genes also showed involvement in response to organophosphorus agents, which are known acetylcholinesterase (AChE) inhibitors, supporting a cholinergic participation.

**Figure 4.**
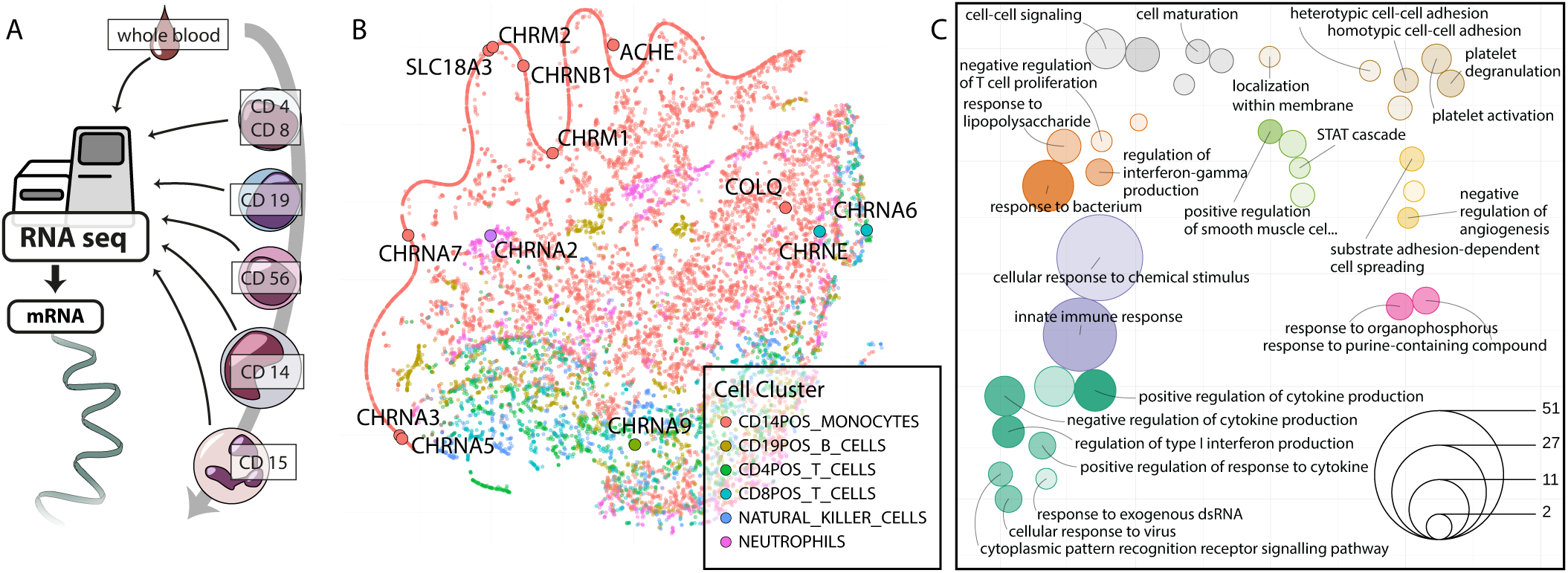
Immune cell gene expression clustering and long RNA pathways perturbed in stroke blood. A) Published cell type-specific long RNA profiles (25) were used to visualize transcriptomes of T lymphocytes (CD4^+^ T-helper cells and CD8^+^ T-cytotoxic cells), B lymphocytes (CD19^+^), NK cells (CD56^+^), monocytes (CD14^+^), and neutrophils (CD15^+^). B) t-SNE visualization of 15032 genes on the basis of their expression in blood-borne immune cells extrapolated from transcriptional activities in regulatory circuits (25). Genes are colored by the cell type in which their expression was highest. Cholinergic core and receptor genes were mainly found in the CD14^+^ monocytic compartment. C) Enrichment of post-stroke DE genes (log2FC > 1.4) in circulation- and immunity-related pathways, presented as t-SNE of GO terms by their shared genes (see Supplementary Methods); color denotes t-SNE cluster, size denotes number of significant genes in term; deeper color indicates lower enrichment p-value (all p-values < 0.05). Distance between terms indicates the number of shared genes between the GO terms, closer meaning more shared genes.

#### Stroke leads to perturbation of microRNA regulatory networks

Notably, miRs upregulated after stroke both appear in smaller numbers compared to downregulated miRs and possess significantly fewer gene targets per individual miR (via *miRNeo*, mean [up vs down] 463 vs 804, median 266 vs 717, one-way ANOVA p = 2.1e-07, F(1,352) = 28.06). To unravel target genes of these two miR populations, we performed gene target enrichment via permutation inside *miRNeo* (10,000 permutations). Significantly enriched gene targets (permutation p-value < 0.05) were subjected to GO analyses and visualized in a t-SNE projection, yielding 13 clusters of related terms, indicating most-affected pathways (Fig. 5A, Fig. S5). Within each cluster, we determined the most relevant genes via hypergeometric enrichment (Fisher’s exact test). Ranking of the clusters by the absolute number of enriched genes (Benjamini-Hochberg adjusted p < 0.05) revealed the putative biological processes that were most influenced by the miR perturbation following ischemic stroke in our patients (Fig. 5B).

**Figure 5.**
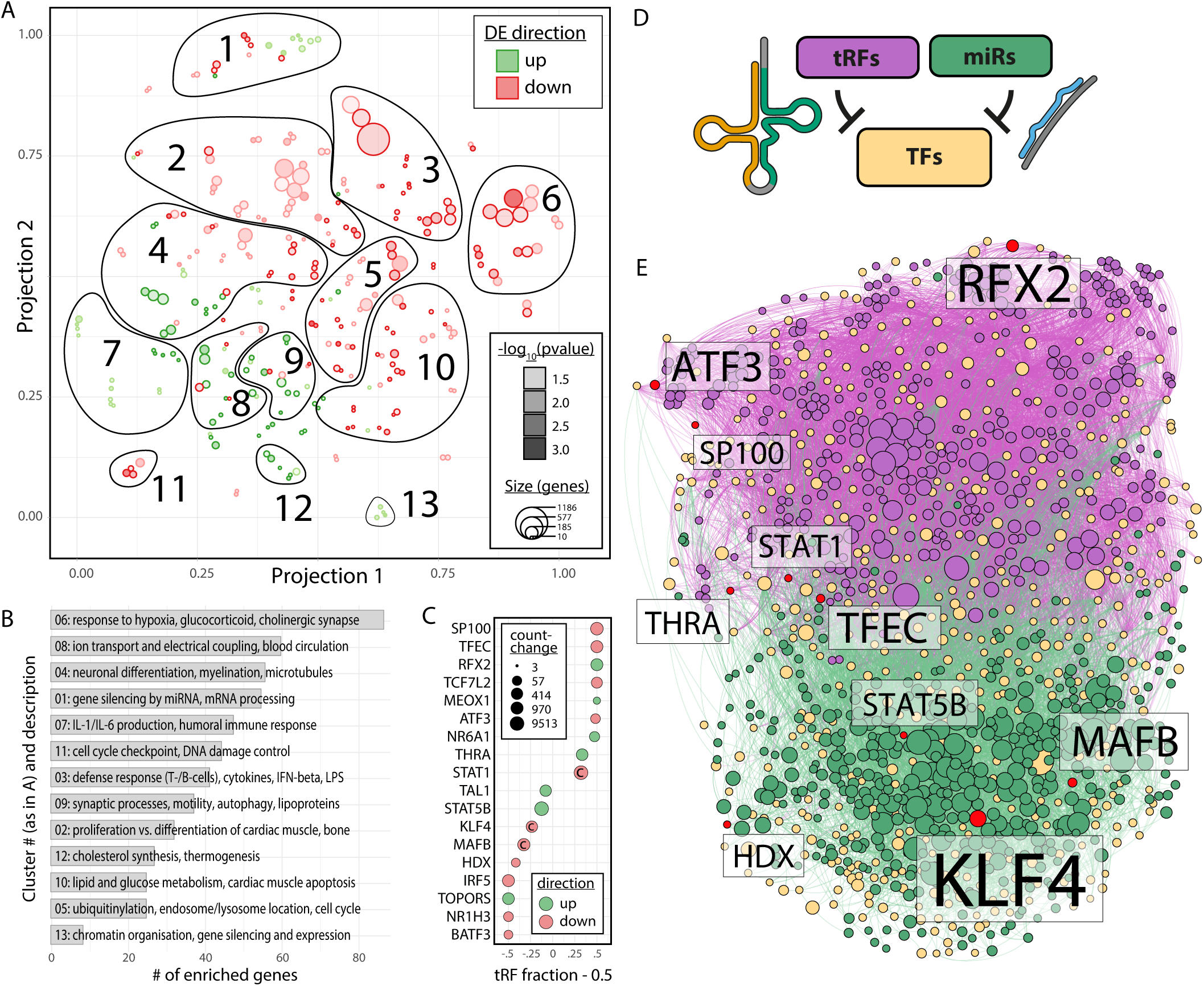
GO enrichment of miR targets and perturbed pathways; divergent influence of miRs and tRFs in CD14^+^ TF regulatory circuits. A) t-SNE visualization of GO terms enriched in the targets of miRs perturbed by stroke, performed separately for positively (green) and negatively (red) perturbed miRs, segregated into 13 functional clusters. Size of circles represents the number of genes in the respective GO term; depth of color represents enrichment p-value (all p < 0.05). B) Bar graph of clusters identified in *A)* ordered by the number of enriched genes (Fisher’s exact test, BH adjusted p < 0.05) shows most pertinent processes with miR involvement. C) The top 18 DE TFs in stroke patients’ blood present a gradient of targeting by miRs and/or tRFs (left = 100% miR targeting, right = 100% tRF targeting; value shown as “tRF fraction – 0.5” to center on 50/50 regulation by miRs and tRFs). Size of points and color denote absolute count-change and direction of differential regulation, respectively. “C” marks TFs targeting cholinergic core or receptor genes. D) Small RNA targeting of TFs active in CD14^+^ cells was analyzed using an in-house database (18). E) Force-directed network of all TFs active in CD14^+^ monocytes self-segregates to form largely distinct TF clusters targeted by DE tRFs and miRs in stroke patients’ blood. Yellow = TF, red = TF DE in stroke patients’ blood, green = miR, purple = tRF. Size of node denotes activity towards targets.

The significantly de-repressed cluster no. 6 (84 enriched genes) pointed towards perturbation of pathways involved in responses to hypoxia (GO:0036293, p = 0.008) and drugs (GO:0042493, p = 0.035), including antibiotics (GO:0071236, p = 0.013), glucocorticoids (GO:0051384, p = 0.007), and the cholinesterase-blocking organophosphorus agents (GO:0046683, p = 0.019), which reinforced the notion of cholinergic participation. Cross-check of enriched genes via DAVID (26) confirmed a role of cluster no. 6 in hypoxia (GO:0071456, p = 8.3e-13), drug response (GO:0042493, 5.4e-10), and the cholinergic synapse (KEGG pathway 04725, p = 0.006). Cluster no. 8 (with second-most 58 enriched genes) highlighted perturbed transmembrane ion conductivity, particularly in regulation of cardiac muscle cell action (GO:0098901, p = 0.044) and negative regulation of blood circulation (GO:1903523, p = 0.016). DAVID analysis confirmed involvement in ‘*regulation of cardiac muscle contraction by the release of sequestered calcium ion*’ (GO:0010881, p = 2.1e-15) and regulation of heart rate (GO:0002027, p = 1.2e-07). The subsequent clusters indicate further involvement in nerve cell regulatory processes (cluster no. 4), regulation of gene silencing by miRNA (cluster no. 1), and humoral immunity via IL-1 and IL-6 (cluster no. 7) (see also Fig. S5). The entire list of clusters and gene enrichments is available as Data File S6.

#### tRFs may suppress inflammation and cholinergic-associated transcription factors alone or in cooperation with miRs

Cellular responses to different stimuli are coordinated by cell type-specific transcriptional regulatory circuits. To facilitate understanding of the role of miRs and tRFs in regulating the transcriptional state of CD14^+^ monocytes after stroke, we generated a monocyte-specific transcriptional interaction network of small RNAs targeting transcription factors (via *miRNeo*) (19), combined with differential expression of long and small RNAs from the PREDICT cohort. The gradually divergent targeting of these TFs by miRs and/or tRFs implied largely separate domains of regulation by these small RNA species (Fig. 5C). This notion was topologically strengthened by the fact that the force-directed network of all TFs active in CD14^+^ monocytes self-segregated to form two distinct clusters of TFs which were primarily targeted either by miRs or tRFs, including numerous TFs DE in stroke patient blood (Fig. 5D&E). Among the implied TFs are proteins known for their influence on cholinergic genes as well as their involvement in inflammation, such as STAT1 or KLF4 (27, 28). Notably, 8 DE TFs were not predicted to be targeted by any miR or tRF present in CD14^+^ monocytes.

#### Stroke-induced tRFs show evolutionarily conserved participation in macrophage/monocyte responses to inflammatory stimuli

To test if the stroke-induced tRFs are involved in the inflammatory response of monocytes and macrophages, we subjected murine RAW 264.7 cells to LPS-stimulation with or without dexamethasone suppression of their inflammatory reactions (Fig. 6A). By 18h after LPS stimulation, RT-qPCR analysis after size-selection (for ≤50 nt fragments) detected pronounced upregulation of the top 6 post-stroke upregulated tRFs (Fig. 6B). Moreover, dexamethasone suppression of the LPS response led to downregulation of those tRFs, along with a diminished inflammatory response (Fig. S6). Predicted targets of these molecules comprise members of mitogen-activated protein kinases (MAPK) and tumor necrosis factor receptor-associated factors (TRAF) (see Data File S7), further pointing towards their regulatory role in response to inflammatory stimuli. Notably, one of the top 6 stroke-perturbed tRFs, tRF-22-WE8SPOX52, is complementary to the 3’UTR sequence of murine Z-DNA binding protein Zbp1 and therefore a predicted regulator of the Zbp1 transcript and its immune system activity (Fig. 6C). To challenge the functional activity of tRF-22-WE8SPOX52 in murine RAW 264.7 macrophages, we over-expressed this tRF using ssRNA mimics (Fig. 6D, Fig S7), which significantly reduced the expression of its Zbp1 target compared to negative control (NC), as quantified by long RNA sequencing (Fig. 6E) and validated by RT-qPCR in an independent experiment (Fig. 6F).

**Figure 6.**
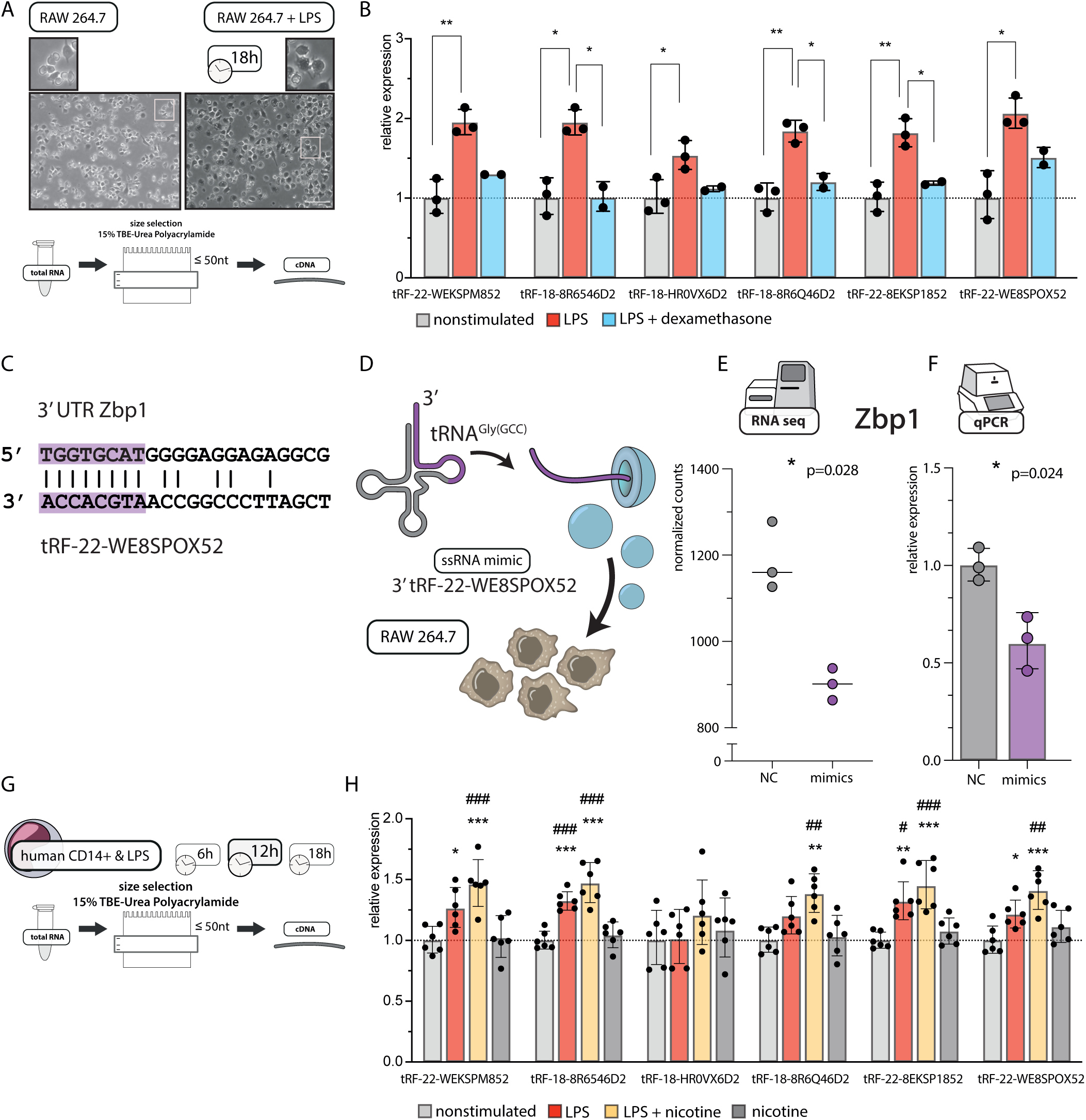
tRF changes upon LPS stimulation of murine RAW 264.7 macrophages and human CD14^+^ monocytes and tRF-mimic transfection. A) LPS stimulation of RAW 264.7 murine macrophage cells induced clear morphologic changes within 18h. Extracted RNA was subjected to size selection and cDNA synthesized from the ≤50 nt fraction alone. Scale bar = 100μm. B) LPS-stimulated RAW 264.7 cells show dexamethasone-suppressible elevated levels of post-stroke induced tRFs. Normalized RT-qPCR values (using mmu-miR-30d-5p, mmu-let7d-5p as house-keeping transcripts, Supplementary Methods), compared to unstimulated controls. Each dot represents 2-4 technical replicates, ANOVA with Tukey post-hoc, * p < 0.05, ** p < 0.01, bar graphs +/- standard deviation (SD(lg)). C) Murine Zbp1 sequence carries an 8 nucleotides-long fragment in the 3’UTR complementary to tRF-22-WE8SPOX52. D) To test the miR-like mechanism of action, RAW 264.7 cells were transfected with ssRNA tRF-22-WE8SPOX mimics or negative control ssRNA, and RNA was extracted 24h after transfection and subjected to polyA-selected RNA sequencing (E) and RT-qPCR (F). E) Long RNA sequencing of cells transfected with ssRNA tRF-22-WE8SPOX52 mimics revealed significantly downregulated expression of Z-DNA binding protein (Zbp1) as compared to negative control (NC). * p < 0.05, shown is adjusted p-value of Wald test via DESeq2, bar graph +/- SD. F) RT-qPCR from an independent cell culture experiment confirmed the downregulation of Zbp1 expression after ssRNA tRF-22-WE8SPOX52 mimic transfection (relative normalized expression using Gapdh as a house-keeping gene), * p < 0.05 one-way ANOVA, each dot represents a technical cell culture replicate, bar graph +/-SD(lg) G) MACS-sorted CD14^+^ cells from healthy human donors were stimulated with LPS with or without addition of nicotine and collected 6h, 12h and 18h thereafter. Extracted RNA was subjected to size selection and cDNA synthesized from the ≤50 nt fraction. Timepoints 6h and 18h are shown in Figure S8. H) At 12h after LPS stimulation, human monocytes exhibited upregulation of post-stroke induced tRFs as compared to unstimulated controls or cells treated with nicotine alone. This reaction was boosted by the addition of nicotine. Shown is relative expression (hsa-miR-30d and hsa-let7d-5p were used as house-keeping transcripts, see Supplementary Methods) normalized to the nonstimulated group. Each dot represents one donor. ANOVA with Tukey post-hoc, * p < 0.05, ** p < 0.01, *** p < 0.001 vs. non-stimulated cells; ^#^ p < 0.05, ^##^ p < 0.01, ^###^ p < 0.001 vs. cells upon addition of nicotine, bar graphs +/- SD(lg).

Lastly, we aimed to validate the functional implications of these findings in primary human cells. Therefore, we performed LPS stimulation experiments in magnetic-activated cell sorted (MACS) CD14^+^ monocytes from healthy volunteers and collected the cells at 6h, 12h and 18h after LPS addition (Fig. 6 G&H, Fig. S8 A&B for 6h and 18h timepoints). To further challenge the cholinergic link, we used nicotine as an immunosuppressive agent (Fig. S8C) (29). LPS-stimulated primary CD14^+^ cells presented significant upregulation of 4 out of the 6 stroke-induced tRFs at 12h, an effect which was augmented by the addition of nicotine. Interestingly, at 18h, the tRF levels in the LPS + nicotine group were comparable to those of nonstimulated cells (Fig. S8B). Together, these findings demonstrate evolutionarily conserved and cholinergic-regulated increases of stroke-induced tRFs under pro-inflammatory insults.

## Discussion

To date, few studies have simultaneously assessed the joint impact of blood miR and tRF-changes in human disease. Here we have discovered a stroke-induced decline of miRs and concomitant elevation of tRFs in whole blood, and demonstrated that this shift may be associated with the post-stroke cholinergic blockade of immune function. To validate our RNA-seq findings of tRNA fragments in a way that circumvents the ambiguous detection of full-length tRNA, we developed and used a size selection-based RT-qPCR test in an independent cohort of patients. Mining transcriptomic datasets identified CD14^+^ monocytes as likely pivotal in the cholinergic control of immunity, demonstrated that the stroke-induced tRFs may target specific monocytic TFs, and showed that at least some of those tRFs may actively control processes linked to inflammatory responses. Moreover, several of the stroke-induced tRFs were also induced in LPS-exposed murine macrophages and in human CD14^+^ primary cells and showed time-dependent nicotine- and dexamethasone-induced upregulation/suppression, supporting the notion that the elevation of tRFs is an evolutionarily conserved response mechanism. Overexpression of tRF-22-WE8SPOX52 using ssRNA mimics led to the downregulation of its Zbp1 target, which is involved in regulating inflammatory responses. This concept of integrated fine-tuning of post-stroke immune responses opens new venues for stroke diagnostics and therapeutics.

The cholinergic anti-inflammatory reflex plays a substantial role in regulating peripheral immune responses after CNS injury, along with the HPA axis and sympathetic signaling (3). Excessive cholinergic responses suppress pulmonary innate immunity, including macrophage and alveolar epithelial cell responses; this may facilitate the development of pneumonia (30), a major factor of non-recovery (31). However, while reduced AChE activities in post-stroke patients’ serum associate with poor survival (32), stroke-induced immunosuppression may be brain-protective (2), calling for caution when considering therapeutic boosting of immune reaction in the periphery to limit infections. Therefore, an in-depth understanding and characterization of the molecular regulators of immune responses and the cholinergic pathway after CNS-injury is of utmost importance at both the system and mechanism levels.

### System-level perturbations associated with ischemic stroke

We identified CD14^+^ monocytes to be the most likely immune cell subpopulation for a transcriptional cholinergic response. Monocytes play established roles in responses to stroke including prolonged monocytosis, deactivation and functional impairment of circulating monocytes/macrophages observed in experimental models (33) and human patients (34). Moreover, stroke leads to overproduction of CD14^++^/CD16^-^ (classical) and CD14^++^/CD16^+^ (intermediate) monocytes with simultaneous decrease in CD14^+^/CD16^++^ (nonclassical) monocytes, which correlates with stroke-associated infection (35). Relatedly, immune cells in general, and monocytes in particular appear to be enriched in specific small RNA species (compare Fig. 3C and (36)), and several of the most highly-perturbed small RNAs are abundantly expressed in monocytes (compare Fig. S4, (37)).

Our in-depth analysis of the pathways targeted by perturbed miRs supports a tie between stroke-induced changes and a cholinergic response. Both the identified clusters as well as the genes enriched in each cluster may be further investigated for identifying the diverse mechanisms involved. A recent whole blood microarray survey identified 15 miRs, 11 of which were replicated in our study, to be suppressed within less than 72 hours in intracerebral hemorrhage patients compared to controls (38). Those miRs pointed towards the same processes we found, including inflammation and humoral immunity via JAK/STAT-activating cytokines, vascular integrity, and the cellular immune response (38).

Stroke is a sudden incident with rapid onset and drastic systemic changes within a short time frame. In the response to such immunologic emergencies, translational control is an important tool (39) That tRFs may be rapidly produced by regulated nuclease cleavage of pre-existing tRNAs in a “burst-like” fashion makes them particularly appropriate for handling acute situations. Recent reports demonstrate production of 3’-tRFs by specific nucleases, and 3’- and 5’-tRNALeu fragments were shown to regulate T cell activation (40). Furthermore, tRFs can perform different molecular roles, including Ago-mediated suppression of target genes carrying complementary sequence motifs (13). At least two of the stroke-induced tRFs upregulated after LPS stimulation show miR-like function: tRF-22-WE8SPOX52 regulates B cell growth via suppressing the expression of Replication Protein A1 (RPA1) (21) and a 17 nucleotides-long analog of tRF-18-HR0VX6D2 limits cancer cell proliferation by impacting the cholinergic-regulating Notch signaling pathway (22). Interestingly, hsa-miR-1260b, identified in our study and by others as perturbed post-stroke (41) differs from tRF-18-HR0VX6D2 by only one nucleotide at position 9 and an additional nucleotide at the 3’-end (Fig. S9), which indicates that hsa-miR-1260b may actually be a tRF (42), and calls for further investigations.

Our study identified two main factors that may lead to an overall decrease in transcript regulation by miRs after stroke. Firstly, the majority of miRs perturbed in our patient collective were downregulated, and secondly, the downregulated miRs possessed significantly more targets than the upregulated ones. For many processes regulated by these miRs, the resultant effect will hence be de-repression of targeted genes (compare Fig. 5A). Additionally, we have identified a stark dichotomy between the target sets of miRs and tRFs, indicating much complementarity and only little cooperative overlap of affected transcripts between those two small RNA species (Fig. 5E). However, these changes may still lead to a homeostatic *functional* cooperation. In summary, the post-stroke “changing of the guards” in the small RNA response may lead to preferential de-repression of miR targets and concomitant repression of tRF targets, and the de-repression of miR targets may be as pivotal for regulating the initial inflammatory response and subsequent peripheral immunosuppression as the tRF elevation we identified.

Kinetically, stroke is characterized by an initial inflammatory response followed by immunosuppression facilitated by, among others, the cholinergic anti-inflammatory reflex (3). Therefore, the time-dependent elevation of tRFs and Cholino-tRFs in particular may offer new mechanisms of homeostatic fine-tuning in response to cerebral ischemia. Further, not only the peripheral but also the central immune response at the site of the injury is of great importance for stroke prognosis. Brain injury triggers activation of microglia and infiltration of peripheral immune cells, including monocyte-derived macrophages, which accumulate at the lesion site 3-7 days after stroke (35). Experimental evidence highlights essential roles of these cells in CNS-repair processes and neuronal protection (43, 44), and our own studies indicate small RNA-regulated cross-talk between neuronal and immunological regulation by JAK/STAT-related mechanisms (19).

Our current study presents tRFs as potential players in regulating the post-stroke inflammatory responses. For example, KLF4, identified as down-regulated in our sequencing dataset, is involved in controlling the macrophage response to LPS (28) and the differentiation of monocytes towards an inflammatory phenotype (45). Therefore, a decrease in miRs targeting this TF may contribute to pro-inflammatory monocytic response observed in the initial phase of stroke. Similarly, MAFB is essential in facilitating the clearance of damage-associated molecular patterns (DAMPs) in the ischemic brain, and, consequentially, limiting the inflammatory response while supporting recovery (46). MAFB de-repression in peripheral immune cells may be a mechanism supporting monocyte infiltration of the brain. Conversely, STAT1 and ATF3 may be preferentially repressed due to their targeting by tRFs. STAT1 is essential in IFN- and IL-6-mediated inflammatory response, and ATF3 is similarly induced by IFNs and contributes to STAT activity via inhibition of STAT-dephosphorylating phosphatases (47, 48). Additionally, ATF3 down-regulates AChE expression during stress (49). Whether these processes contribute to body homeostasis after the damaging event, or rather to pathologic derailment of immune function, requires detailed kinetic studies of circulating monocytes and brain-infiltrating monocyte-derived macrophages, with simultaneous profiling of short and long transcripts.

### Mechanistic implications of tRF regulation after ischemic stroke

To gain new insight into the regulation of inflammation by stroke-induced tRFs, we quantified the top 6 perturbed tRFs in RAW 264.7 murine macrophages and primary LPS-stimulated CD14^+^ human monocytes. Both cell types responded to LPS by upregulation of these tRFs within 12-18h. Interestingly, dexamethasone prevented or subsequently downregulated the increased expression of tRFs in the RAW 264.7 cells. To further seek cholinergic links of these stroke-regulated tRFs, we used nicotine as an immunosuppressive stimulus in LPS-stimulated human CD14^+^ cells. Monocytes and macrophages express the cholinergic nicotinic alpha 7 receptor, which after binding of acetylcholine downregulates the production of inflammatory cytokines (e.g. TNFα) (29). In human CD14^+^ cells, the levels of the top 6 stroke-induced tRFs were transiently elevated by the addition of nicotine at the 12h timepoint (back to baseline by 18h post-stimulation). Thus, the elevated levels of blood tRFs two days after ischemic stroke may reflect potentiated cholinergic signaling, which remains to be investigated in the clinical setting.

In human patients, the stroke response in blood is cell type- and time-specific. For example, day two post-ischemia features a transient increase in STAT3 phosphorylation of monocyte subsets, which is also detected in patients after major surgery (50). Conversely, STAT3 signaling causes immune stimulation in monocytes but is linked to immunosuppression in monocytic myeloid-derived suppressor cells (M-MDSC) (50). Therefore, the biological activities of stroke-induced tRFs are very likely also cell type- and context-specific. Our ssRNA tRF-22-WE8SPOX52 mimic experiments further support tRF involvement in the posttranscriptional regulation of genes implicated in inflammatory responses. Zbp1, which was significantly downregulated under tRF-22-WE8SPOX52 overexpression, is a DAMP-sensor which induces interferon responses, programmed cell death, and NLRP3 inflammasome formation (51). The ZBP1 protein has often been linked to the response to viral infections, but some studies point to its role in the reaction to bacterial pathogens, where it may be involved in the induction of necroptosis (51). Incidentally, the ZBP1 transcript is also downregulated in our long RNA sequencing of patient blood (logFC = −1.7, adjusted p-value = 0.001); the exact nature of the interaction of tRF-22-WE8SPOX52 and ZBP1 should be subject of future studies.

### Limitations

We hypothesized that the tRFs identified in our study are of cellular origin and therefore re-analyzed the small RNA sequencing data provided by Juzenas et al. (GSE100467). While some studies identified an enrichment of tRFs in exosomes (40), our re-analysis found the tRFs mostly in the cellular blood compartments. Although the main immune populations are included in the analyzed dataset, it should be noted that the tRFs identified in our study may also originate from immune cells not sorted/sequenced by Juzenas et al. Additionally, considering the different roles of specific immune subpopulations, identifying the specific source of tRFs and their roles in immune function should be the goal of further investigations. For example, monocyte subsets are differentially regulated after stroke, and since these cells all express the CD14 marker (35), a higher cellular resolution is called for. Also, tRFs are induced upon cellular stress (52), such that the sorting procedure may affect their expression, and they may reside in extracellular compartments (15, 53), calling for testing the impact of sample processing, RNA isolation and sequencing techniques on the detectability of tRFs. Additionally, a method for the direct comparison of miR- and tRF-levels in the same sequencing experiment will be important for discerning the true difference in detected counts after alignment, possibly via spike-in procedures. The currently maturing technology of single-cell sequencing is an obvious candidate for achieving the goals of higher cellular resolution along with avoiding stressors associated with sample preparation, but its shallow sequencing depth is still an issue.

Given the sex-related differences in cholinergic responses (54), the molecular regulators of cholinergic signaling and immunity should be investigated in detail in both males and females. However, to increase the consistency of our results, we only included male stroke patients, which is a further limitation of this study. Interestingly, the overall impact of stroke is greater in women, as their higher life expectancy is linked to increased stroke incidence in older adults, and they face worse recovery prospects (55). Last, but not least, tRFs may have functions other than their miR-like activities; for instance, tRNALeu-CAG fragments facilitate translation and ribosome biogenesis (12), whereas tRNAGly, tRNAGlu, tRNAAsp and tRNATyr-derived tRFs displace RNA-binding proteins leading to mRNA destabilization (11). Therefore, potential functions of stroke-perturbed tRFs other than Ago-mediated suppression of translation should be further examined.

### Conclusion

While the specific roles of tRFs in regulating local neuroinflammatory responses and functional modulation of specific peripheral monocyte subsets remain to be elucidated, our findings point towards tRFs/miRs as homeostatic regulators of post-stroke immune responses and potential biomarkers for increased infection risk in these patients. The cumulative role of tRFs and miRs as general post-damage mediators of CNS-immune communication thus calls for seeking small RNAs, and tRFs in particular, as involved in other traumatic pathologies such as spinal cord injury, traumatic brain injury, concussion, as well as neuroinflammatory brain diseases.

## Materials and Methods

Expanded methods can be found in the online supplement.

### Clinical cohort

PREDICT was a prospective multi-center study with sites in Germany and Spain (www.clinicaltrials.gov, NCT01079728)(16) that analyzed 484 acute ischemic stroke patients. Patients underwent daily screenings for stroke-associated pneumonia, dysphagia and inflammation markers and their clinical outcome was recorded 3 months post-stroke. To exclude very severe cases of stroke, we only considered for sequencing samples from patients with modified Rankin Scale (mRS) values of 3 and below at discharge from the hospital, leaving n=240 relevant cases. Blood was collected into RNA stabilizing tubes (Tempus Blood RNA tubes, Applied Biosystems™) on each day of hospitalization. Blood samples collected on the 2^nd^ day were subjected to small and long RNA-sequencing, with time from stroke occurrence to blood withdrawal varying between 0.94 to 2.63 days (average: 1.98 days). Blood samples from age- and ethnicity-matched healthy controls were obtained at matched circadian time from donors with ethical approvals from institutional review boards (ZenBio, North Carolina, USA).

### RNA extraction, quality control and sequencing

RNA was extracted from 3 ml of whole blood of 484 PREDICT patients using Tempus Spin RNA isolation kit (Invitrogen, Thermo Fisher Scientific, Waltham MA, USA). Pre-sequencing Bioanalyzer 6000 (Agilent, Santa Clara CA, USA) tests showed high RNA quality (RIN values 7.9-9.9, median 8.8). Libraries constructed from 600 ng total RNA of 43 samples were subjected to small RNA-sequencing (NEBNext® Multiplex Small RNA library prep set for Illumina, New England Biolabs, Ipswich MA, USA), and 24 of the small RNA-sequenced samples served for PolyA-selected long mRNA sequencing (1000 ng total RNA per sample, TruSeq RNA library preparation kit (Illumina, San Diego CA, USA)). Sequencing (24 or 12 samples per flow cell for small and long RNAs, respectively) was performed on the Illumina NextSeq 500 platform at the Hebrew University’s Center for Genomic Technologies.

### Alignment and count table generation of RNA sequencing reads

Quality control was performed using FastQC, version 0.11.2 (56) more details can be found in the Supplementary Methods. Flexbar (with parameters “-q TAIL -qf Sanger -qw 4 -min-read-length 16”) (57) served for adapter trimming and quality based filtering of all raw reads. Long RNA was aligned to the human reference transcriptome (ENSEMBL GRCh38 release 79) using salmon (58) with default parameters. Small RNA was aligned to the miRBase version 21 using miRExpress 2.1.4 (59) with default parameters but skipping adapter trimming for miR expression, and to the tRNA transcriptome using the MINTmap pipeline (60) with default parameters for tRF expression (using only reads mapping exclusively to the tRNA space). Raw gene-expression data of small and long RNA sequencing and technical covariates of all experiments are available via the NCBI GEO database (accession number GSE158312).

### Size selection for tRF quantification

Standard RT-qPCR methods do not allow to distinguish between full length tRNA molecules and 3’tRFs. To exclude longer RNA species in the RT-qPCR quantifications, we performed RNA size selection on 15% TBE-Urea-Polyacrylamide gels, selecting only RNA molecules ≤ 25 nucleotides for validations in the clinical cohort and ≤ 50 nucleotides for the assessment of tRF expression in LPS-stimulated RAW 264.7 cells and human CD14^+^ monocytes. Detailed description can be found in the Supplementary Methods.

### Analysis of the presence/absence of specific tRFs in blood compartments

In descriptive analysis of small RNA expression, a threshold (e.g., at least 5 counts in at least 80% of samples) is often used to define presence or absence of small RNAs. However, since this definition relies heavily on sequencing depth, and depth can vary widely even in methodically robust sequencing experiments depending on a large number of variables, we defined our own test for descriptive analysis of presence or absence of lowly expressed small RNAs in each of the sample types. Briefly, this definition comprises estimation of a log-normal distribution on the expression profile of the small RNA across all samples in the individual cell types, and a statistical test to refute the null hypothesis that the distribution is in fact log-normal. For each small RNA, the distribution mean and standard deviation of the expression values per cell type were estimated using the *fitdist* function of the R/fitdistrplus package (61). The count distribution was then tested against a log-normal distribution with the estimated mean and standard deviation via the R implementation of the Kolmogorov-Smirnov test, with a cutoff of 0.1. The small RNA was defined as present if the test failed to reject the null hypothesis (see Fig. S4 for examples). The code implementation is available at https://github.com/slobentanzer/stroke-trf.

### LPS stimulation of murine macrophages

Murine RAW 264.7 cells (ATCC TIB-71) cultured in Dulbecco’s Modified Eagle’s Medium supplemented with 10% Fetal Calf Serum, 1% Penicillin-Streptomycin-Amphotericin B and 1% L-Glutamine (all reagents from Biological Industries, Beit HaEmek, Israel) were collected using a cell scraper and stimulated with lipopolysaccharide (LPS from *E*.*coli* 0127:B8, Sigma Aldrich, St. Louis, USA) following modified protocol (62). Briefly, 2e05 cells were stimulated with 100 ng/ml LPS +/- 0.5 μM dexamethasone per well (Sigma Aldrich, St. Louis, USA) in 12-well cell culture plates. Cells were collected in Tri-Reagent (Sigma Aldrich, St. Louis, USA) 18h after LPS stimulation and RNA was isolated using miRNeasy kit (Qiagen, Hilden, Germany). For the size selection, 1 μg of total RNA was used and cDNA was synthesized from 500 pg of size-selected RNA using qScript microRNA cDNA Synthesis Kit (Quanta Biosciences, Beverly MA, USA) and following standard protocol (for further details see Supplementary Methods). Data presented in the manuscript is derived from 3 independent experiments (2 of them with dexamethasone treatment) with 2-4 technical replicates in each group.

### Transfection experiments with tRF-22-WE8SPOX52 mimics, RNA sequencing and RT-qPCR

Transfections were performed using HiPerFect transfection agent (Qiagen, Hilden, Germany) following a standard protocol for transfecting RAW 264.7 macrophages. Briefly, 2e05 RAW 264.7 cells per well were seeded in 24-well plates and transfected using 6 μl HiPerFect reagent per well and 50 nM final ssRNA tRF-mimics: /5PHOS/rArU rCrCrC rArCrC rGrCrU rGrCrC rAmCmC rA, using NC5 /5PHOS/rGrC rGrArC rUrArU rArCrG rCrGrC rArArU mAmUrG (both from IDT, Coralville IA, USA) as negative control. Cells were collected after 24h following the transfection in Tri-Reagent (Sigma Aldrich, St. Louis, USA) and total RNA isolated using the miRNeasy kit (Qiagen, Hilden, Germany). Two separate experiments were performed: [1] 180 ng RNA was subjected to long RNA sequencing (KAPA stranded mRNA-seq kit, Roche, Basel Switzerland) and [2] Zbp1 levels were quantified by RT-qPCR (relative expression normalized to Gapdh) after cDNA synthesis (qScript Kit; Quanta Biosciences, Beverly MA, USA) from 100 ng RNA.

### Isolation and *ex vivo* stimulation of human monocytes

This study was approved by the ethics committees of the Charité Universitätsmedizin Berlin (MG Cohort: EA1/281/10). Peripheral blood mononuclear cells (PBMCs) were separated from whole blood anticoagulated with heparin by density gradient centrifugation over Ficoll (Biocoll separating solution, Biochrom GmbH, Berlin, Germany). Untouched monocytes were isolated by using a commercially available Pan Monocyte Isolation Kit (Miltenyi Biotec, Bergisch Gladbach, Germany). Cells (2e06 cells/ml) were cultured in RPMI 1640 Medium (VWR, Radnor, USA), supplemented with 1% penicillin-streptomycin (Biochrom GmbH, Berlin, Germany), 2mM L-Glutamine (Biochrom GmbH, Berlin, Germany) and 10% autologous serum and stimulated with LPS (1ng/ml, 0127:B8, Sigma, Kawasaki, Japan) in the presence or absence of nicotine (300µM, Sigma, Kawasaki, Japan) for 6h, 12h and 18h at 37°C. Unstimulated monocytes and monocytes stimulated with nicotine served as controls. Tumor necrosis factor α (TNFα) concentration was measured in cell culture supernatant by using a commercially available DuoSet ELISA kit (R&D systems, Minneapolis, USA). Cells were collected in Tri-Reagent (Sigma Aldrich, St. Louis, USA) and RNA was isolated using miRNeasy kit (Qiagen, Hilden, Germany). For the size selection, 600 ng of total RNA (or maximum loading volume of 20 μl) were used and cDNA was synthesized from 500 pg of size-selected RNA using qScript microRNA cDNA Synthesis Kit (Quanta Biosciences, Beverly MA, USA) and following standard protocol (for further details see Supplementary Methods).

### Statistical analysis

Data analysis was performed using R (version 4.0.2), the code is available at https://github.com/slobentanzer/stroke-trf (including code for analyses of RT-qPCR data) FDR correction was applied whenever applicable. RT-qPCR data was analyzed using Bio-Rad CFX Maestro software (Bio-Rad, Version 4.1.2433.1219) and plotted in GraphPad Prism 8.0 (GraphPad Prism Software Inc., San Diego, USA).

## Supporting information

Main Supplement File

## Data Availability

Sequencing data are available at NCBI GEO: GSE100467.
Code used in analyses is available at https://github.com/slobentanzer/stroke-trf.

## Acknowledgments

The authors would like to thank Dr. Simonas Juzenas (Saarbrücken) and Prof. Andreas Keller (Saarbrücken/Stanford) for their support concerning the blood compartments RNA-sequencing dataset and Dr. Iftach Shaked (San Diego) for fruitful discussions.

