## Supplementary material for "Transfer RNA fragments replace microRNA regulators of the cholinergic post-stroke immune blockade": Main Supplement File

*Expanded Methods:*

**Sequencing Quality Control**
Small RNA was aligned using miRExpress and MINTmap. Total reads (after flexbar with quality control): 500,488,798. Reads per sample: 10,214,057 +/- (SD) 4,605,190. No significant difference between groups. Total miRNA: 284,749,858. miRNA hit / (hit+nohit): 56% +/- (SD) 4.4%. Total tRF (exclusive to tRNA space, see MINTmap documentation): 1,637,476. tRF (exclusive to tRNA space, see MINTmap documentation) hit / total reads: 3238 ppm +/- (SD) 1421 ppm. FastQC: All analyzed samples/bases showed exceptional quality. These metrics and FastQC results can be viewed at https://github.com/slobentanzer/stroke-trf.

At present, no standardized method exists to directly compare absolute counts derived from two separate alignments of the same sequencing experiment, as is the case here with the miRExpress and MINTmap alignments; MINTmap is a much more stringent pipeline (at least for the "exclusive tRNA space" used here). Future experiments should include a spike-in procedure or a similar approach to estimate absolute transcript numbers for each smRNA species, and to enable a better comparison between those absolute values.

Large RNA: Total reads (after flexbar with quality control): 880,069,396. Reads per sample: 36,669,558 +/-(SD) 4,543,364. No significant difference between groups. Mean mapping rate 89% +/- (SD) 2%. FastQC: All analyzed samples/bases showed exceptional quality. These metrics and FastQC results can be viewed at https://github.com/slobentanzer/stroke-trf.

**Differential expression analysis**
Differential expression was determined using the R/DESeq2 package (1) including the log2 fold change shrinkage estimation "apeglm" algorithm (2). Genes were considered differentially expressed at an adjusted p-value of < 0.05. The analysis was performed on the raw count tables (outliers already removed) with correction of covariates (age, batch, time2blood) in the model formula. Outliers were identified by sample clustering based on batch-corrected variance-stabilized expression, leading to exclusion of sample 4044 (stroke patient) in the small RNA sequencing experiment.

**The count-change metric**
The log-fold change metric is not ideally suited for assessing the potential impact of expression changes for individual small RNAs, because it does not reflect mean expression levels. We calculated the count-change for individual miRs and tRFs by combining base mean expression with the de-logarithmicized fold-change (from DESeq2 output).

$$CC = (BM \times 2^{LFC}) - BM$$

CC: countChange, BM: baseMean, LFC: log2-fold change

**Small RNA targeting predictions**
miR targeting was analyzed via our in-house database, *miRNeo*, as described (3). Briefly, the database unites 10 prediction algorithms and all available experimentally validated miR:gene interactions, and unites them in a scoring system. For all analyses in this manuscript, the cutoff score for consideration of a valid miR:gene interaction was ≥ 6.

Since no comprehensive tRF targeting predictions are available, we performed our own prediction based on all tRFs detected with more than 10 reads on average. We used the TargetScan 7.0 algorithm (4) to determine putative miR-like binding of any 7-nucleotide substring ("seed") of any tRF to any human transcript 3'-UTR. Hits were scored according to conserved branch length (BL) and probability of conserved targeting (PCT) across all 23 available species (5) and were entered into *miRNeo* (seed-gene targeting and tRF-seed association).

**Gene set compilation of cholinergic and associated genes**
We started out with a set of 28 cholinergic genes described in (6), targets including ACLY, CHAT, VAChT (aka SLC18A3), the 16 nicotinic and 5 muscarinic receptor subtypes, ACHE, BCHE, PRIMA1, and CHT (aka SLC5A7). In addition, we added several groups of genes known to be associated with

cholinergic functioning and of interest to our aims. These include: genes from the neurokine and neurotrophin signaling pathways; families of genes implicated in the development and differentiation of cholinergic cell types, including lim-homeobox transcription factors, bone morphogenetic protein family genes, and genes from the JAK-STAT pathway (see Data File S2)

miRs were considered Cholino-miRs if they were validated or predicted (with a score > 5/10) to target at least 5 transcripts on this list; tRFs were considered Cholino-tRFs if they contained a seed targeting evolutionarily conserved binding sites in at least 5 transcripts on this list (Figure S2 for empirical cumulative distribution function, ECDF plot); because of diverging numbers between miRs and tRFs we chose to use the higher (more stringent) threshold for miRs. Transcription factors were similarly associated with each transcript on the list, using the transcription factor activity derived from (7) in circulatory immune cells.

**Permutation targeting analysis**
Where appropriate, we determined permutation p-values via *miRNeo* targeting permutation. We determined a score for the test condition (e.g. by summing the individual targeting scores of all DE miRs towards cholinergic genes) and compared it to a null distribution consisting of permutated scores resulting from random substitution of test parameters (e.g. a random selection out of all genes the same size as the cholinergic test set), the p-value being the fraction of the null distribution at least as extreme as the real score.

**Gene Ontology Analyses**
We performed GO analyses on differentially expressed (DE) long RNA transcripts based on their DE p-value. GO analyses were performed using R/topGO as recommended by the authors, using the weighted method (8). Transcripts were ranked by p-value, and all DE transcripts (adjusted p-value < 0.05) with absolute log2 fold changes > 1.4 were tested against the background of the topmost two thousand transcripts. While GO enrichment analysis can be informative, interpretation and visualization of its results is not standardized and is often limited to presentation of top *X* terms by p-value. R/gsoap (9) is an analysis tool proposed to aid in interpretation of GO enrichment results via t-SNE display of similarity of terms based on the amount of shared significant genes. GO enrichment results were processed to fulfil gsoap input criteria and visualized using R/ggplot2 (10).

**Pathways targeted by perturbed miRs**
We determined target sets of positively and negatively DE miRs (separately) via *miRNeo* query with a threshold score of at least 6. The gene targets were ranked by the amount of miRs targeting each gene, yielding a range of 1-44 (mean 4.7 +/-SD 4.7) for positively regulated miRs, and a range of 1-155 (mean 13.5 +/-SD 15.6) for negatively regulated miRs, both resembling power law distributions. To account for possible biases in the underlying targeting dataset, targeting and scoring were then permutated for both sets with the same query and ranking, but randomized input miRs were selected from all mature miRs in the same length as the original DE set, for 10,000 times. Upon calculation of permutation p-values, we dismissed genes that were not targeted by the set of DE miRs significantly more than by the random permutation sets ($p < 0.05$). The larger negative-DE set in this analysis yielded a large number of highly enriched genes, such that a distinction by adjusted p-value was not possible in the top 2000 targeted genes. Thus, we repeated the analysis for negatively regulated miRs using a log2FoldChange threshold of -2, which reduced the amount of miRs to 82, and the range of miRs targeting each gene to 1-52, mean 5.8 +/-SD 6.1. This set was subjected to the same permutation approach and was used instead of the non-limited set in the p-value-based approach below.

The gene lists so derived were used in a GO analysis similar to the one described above, based on the following scorings: for positively regulated miRs, GO enrichment was based on *1)* the significantly targeted genes ($p < 0.05$) as a test set, and the (non-enriched, $p > 0.05$) genes as background; and *2)*, to identify the most-targeted genes, the top 200 genes (sorted by amount of distinct miRs targeting each gene (also $p < 0.05$)) served as a test set, with all other genes (starting from 201) as the background. For negatively regulated miRs, target genes were analyzed similarly, but the non-threshold significantly targeted genes were replaced by the -2 log2FoldChange threshold set in approach *1)*. The GO terms yielded by these four analyses were visualized and clustered using the R/gsoap-approach described above.

The resulting 13 clusters were manually annotated and organized on a canvas to better understand the relationships between the targeted gene sets of positively and negatively regulated miRs (Figure S5).

Few terms fell outside the area of these manually adjoined clusters due to little similarity with the clustered terms; these were disregarded in the further analyses (but are available in Data File S6). To discern the most relevant genes and seek putative hub genes in the stroke response, the clusters identified in the gsoap visualization were imported back into the R environment. Detection of significantly enriched genes in each cluster involved hypergeometric enrichment of each gene in each cluster versus all other clusters (background) via Fisher's exact test. The resultant p-values were corrected for multiple testing using the Benjamini-Hochberg method, and genes with an adjusted p-value < 0.05 were considered significantly enriched in the respective cluster (Data File S6), with a range of enriched genes per cluster from 8 to 84. The final 13 clusters were ranked by the total amount of enriched genes per cluster, and cross-checked for functional implications of enriched genes in each cluster using DAVID 6.8 (11).

**PCR quantification of inflammatory markers after LPS stimulation and Zbp1 quantification.**
**LPS stimulation:** 18h after LPS stimulation, cells were collected in Tri-Reagent (Sigma Aldrich, St. Louis, USA) and total RNA, including small RNAs, isolated using miRNeasy (Qiagen, Hilden, Germany). cDNA was synthesized from 100ng RNA using qScript™ cDNA Synthesis Kit (Quanta Biosciences, Beverly MA, USA). Expression of Cd14, Stat1, Tnfa and Il10 were assessed using PerfeCTa® SYBR® Green FastMix® (Quanta Biosciences, Beverly MA, USA) and normalized using Peptidylprolyl isomerase A (Ppia) and 18S ribosomal RNA genes as house-keeping.

**ssRNA tRF-22-WE8SPOX52 mimics experiments:** 24h after transfection of the mimics, cells were collected in TRI-Reagent (Sigma Aldrich, St. Louis, USA) and total RNA, including small RNAs, isolated using miRNeasy (Qiagen, Hilden, Germany). cDNA was synthesized from 100ng RNA using qScript™ cDNA Synthesis Kit (Quanta Biosciences, Beverly MA, USA), and expression of Zbp1 was assessed using PerfeCTa® SYBR® Green FastMix® (Quanta Biosciences, Beverly MA, USA) and normalized using Glyceraldehyde 3-phosphate dehydrogenase (Gapdh) as house-keeping. Primer sequences were obtained from the Primer Bank (12) and were as follows:

Cd14: **FORWARD** AGCACACTCGCTCAACTTTTC,
**REVERSE** GCCCAATTCAGGATTGTCAGAC
Stat1:**F** TCACAGTGGTTCGAGCTTCAG, **R** CGAGACATCATAGGCAGCGTG
Tnfa: **F** CCTGTAGCCCACGTCGTAG, **R** GGGAGTAGACAAGGTACAACCC
Il-10: **F** GCTCTTACTGACTGGCATGAG, **R** CGCAGCTCTAGGAGCATGTG
Ppia: **F** GAGCTGTTTGCAGACAAAGTTC, **R** CCCTGGCACATGAATCCTGG
18S rRNA: **F** CTCAACACGGGAAACCTCAC, **R** CGCTCCACCAACTAAGAACG
Gapdh: **F** ATCAAGAAGGTGGTGAA, **R** CTTACTCCTTGGAGGCCAT

**PCR quantification of tRFs**
First, we performed size selection of total RNA, to exclude molecules > 25 nt in case of human samples and > 50 nt in case of cell culture experiments with RAW 264.7 cells. 1 μg of RNA was loaded into 15% TBE-Urea-Polyacrylamide gel (Biorad, Hercules, USA) after mixing 1:1 with Gel Loading Buffer II (Thermo Fisher, Waltham, USA) and run at 200V for 40-50 minutes. When testing human CD14+ monocytes, 600 ng of RNA were loaded into the gel; in case of low total RNA concentrations (with minimum 21 ng/μl), we used the maximum accommodable volume of 20 μl. Gels were stained with SYBR Gold (Thermo Fisher, Waltham, USA) and visualized on a UV table to cut out the desired section. As size markers, miR marker and Low Range ssRNA ladder (both from New England Biolabs, Ipswitch, USA) were used. Excised gel fragments were incubated in 810μl 3M NaCl over-night at 4°C on a rotation stand. The supernatant was then transferred into a new Eppendorf and 1 volume of iso-propanol was added for 24h at -20°C. Then, RNA was precipitated.

RNA concentrations were measured using Bioanalyzer (Agilent, Santa Clara, USAμ) and cDNA was prepared from 500 pg (with exception of control samples 16 and 18 yielding low concentrations after size selection – here 250pg were used) using qScript™ microRNA cDNA Synthesis Kit (Quanta Biosciences, Beverly MA, USA) and diluted to 200 μl total. tRFs were quantified using quantitative reverse transcription PCR (RT-qPCR) with PerfeCTa® SYBR® Green FastMix®, Low ROX™ (Quanta Biosciences, Beverly MA, USA) and normalized to hsa-miR-30d-5p, hsa-let-7d-5p, hsa-miR-106b-3p and hsa-miR-3615 (human stroke patients) or mmu-miR-30d-5p, mmu-let-7d-5p for experiments with RAW 264.7 cells. In experiments with ssRNA tRF-22-WE8SPOX52 mimics, tRF-22-WE8SPOX52 levels were assessed without size-selection (measuring levels of the tRF and its parental tRNA molecule) and normalized to Snord47.

Primer sequences are listed below:
hsa/mmu-miR-30d-5p: TGTAAACATCCCCGACTGGAAG
hsa/mmu-let-7d-5p: AGAGGTAGTAGGTTGCATAGTT
hsa-miR-106b-3p: CCGCACTGTGGGTACTTGCTGC
hsa-miR-3615: TCTCTCGGCTCCTCGCGGCTC
tRF-22-WEKSPM852: TCGATCCCCGGCATCTCCACCA
tRF-22-WE8SPOX52 (and tRF-21-WE8SPOX5D): TCGATTCCCGGCCAATGCACC
tRF-18-8R6Q46D2: TCCCCGGCATCTCCACCA
tRF-22-8EKSP1852: TCAATCCCCGGCACCTCCACCA
tRF-18-8R6546D2: TCCCCGGCACCTCCACCA
tRF-18-HR0VX6D2: ATCCCACCGCTGCCACCA
Snord47 (Quanta Biosciences, Beverly MA, USA):
GTGATGATTCTGCCAAATGATACAAAGTGATATCACCTTTAAACCGTTCCATTTTATTTCTGAGG

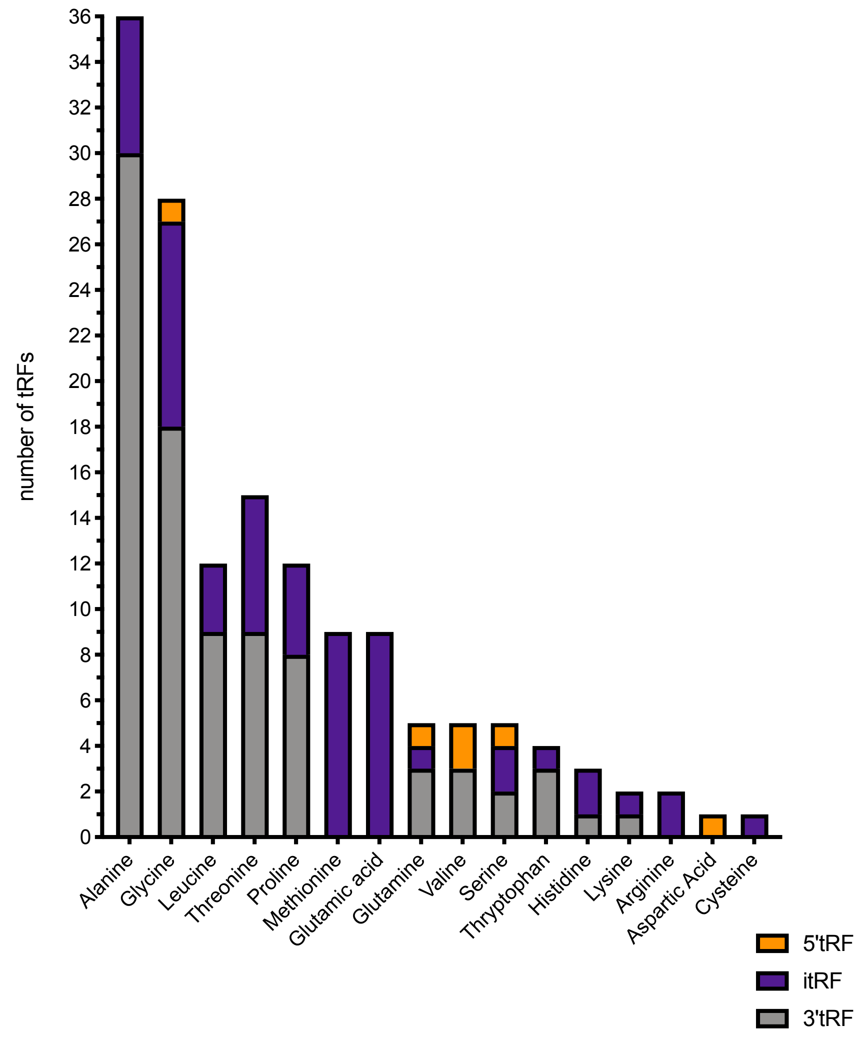


**Figure S1**

**Analysis of types of DE tRFs indicates non-random cleavage of tRNA molecules.**

Most DE tRFs were derived from tRNA-Ala (36) and 3’-tRFs were the most common type in the whole DE dataset.

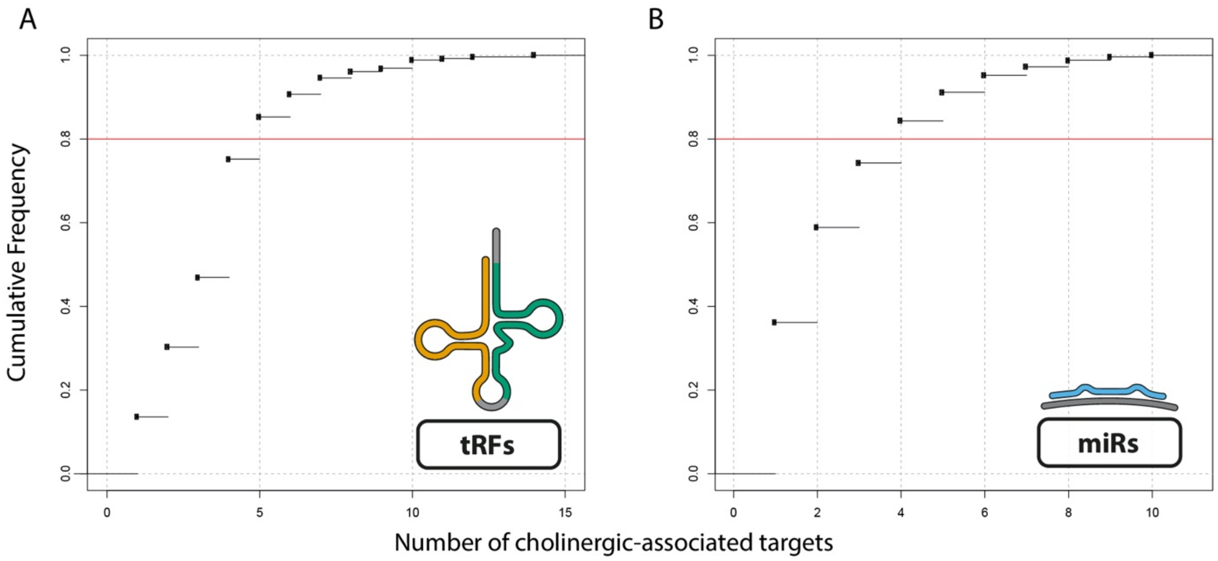


**Figure S2**

**Cholinergic-associated small RNA empirical cumulative distribution function (ECDF) curves.**
Cholinergic association was tested using *miRNeo* targeting data of miRs and tRFs.

To assess the best-suited threshold for defining cholinergic association, ECDFs were calculated for the number of cholinergic-associated (CA) genes targeted by each unique small RNA. A) Cumulative frequency of the number of CA genes targeted by tRFs. Threshold of 80% (red line) is passed at five CA genes targeted. B) Cumulative frequency of the number of CA genes targeted by miRs. Threshold of 80% (red line) is passed at four CA genes targeted.

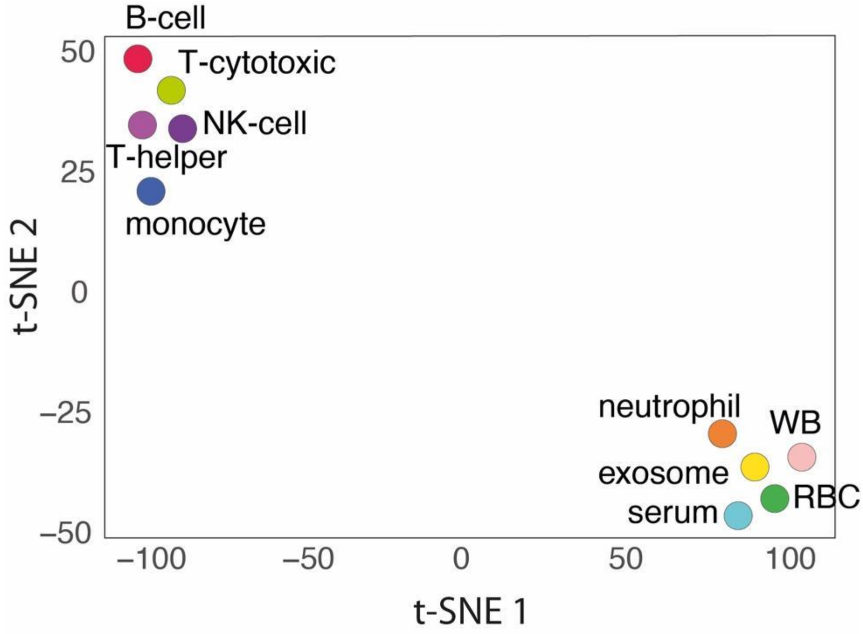


**Figure S3**

**tRF profiles of blood compartments distinguish leukocyte subsets, excluding neutrophils, from erythrocytes and non-cellular compartments**. t-SNE of specific blood compartments based on tRF expression, WB – whole blood, RBC – erythrocytes.

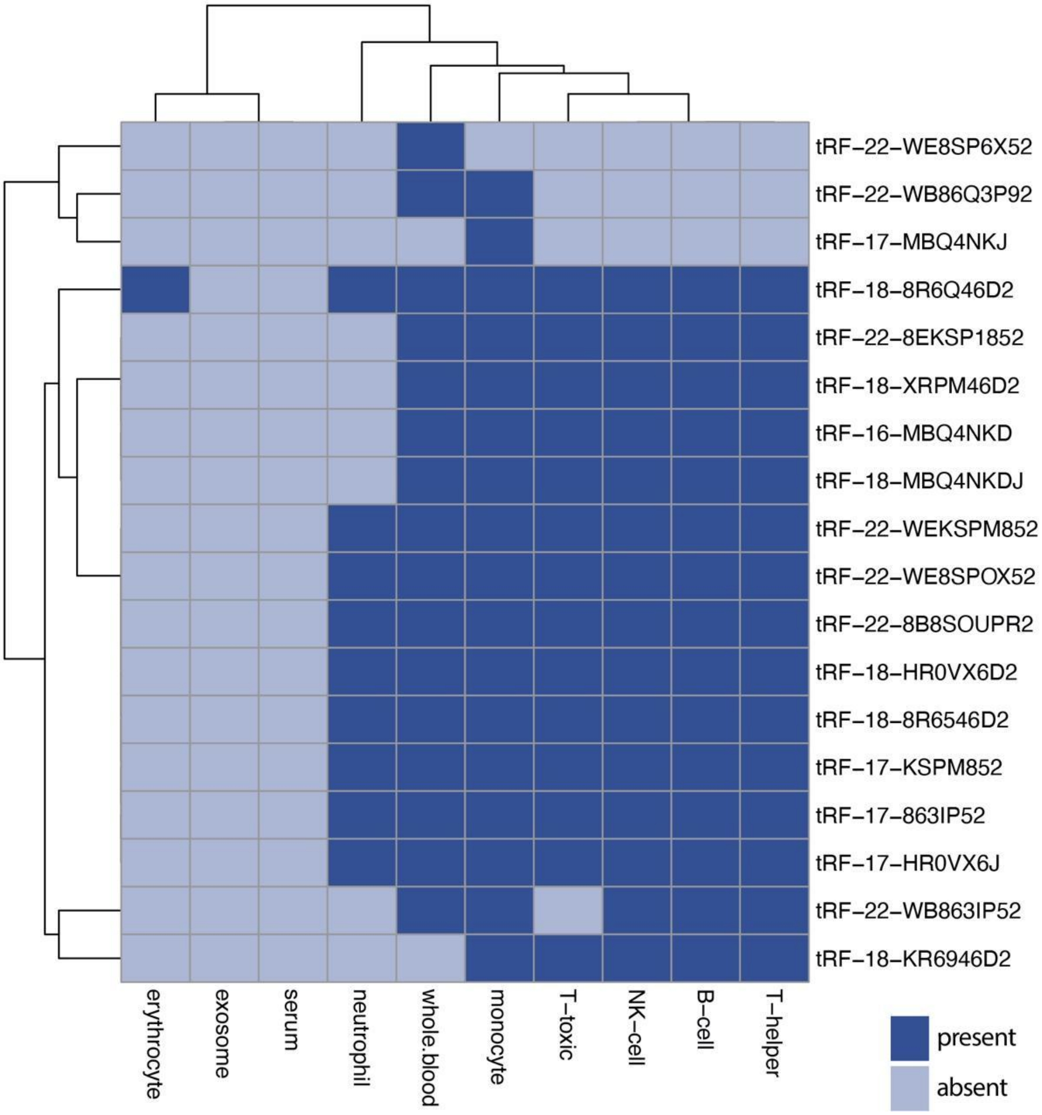


**Figure S4**
**Most of top 20 stroke DE tRFs are present in immune cell compartments.** Heatmap showing the presence/absence of the 20 most highly-perturbed tRFs in defined blood compartments.

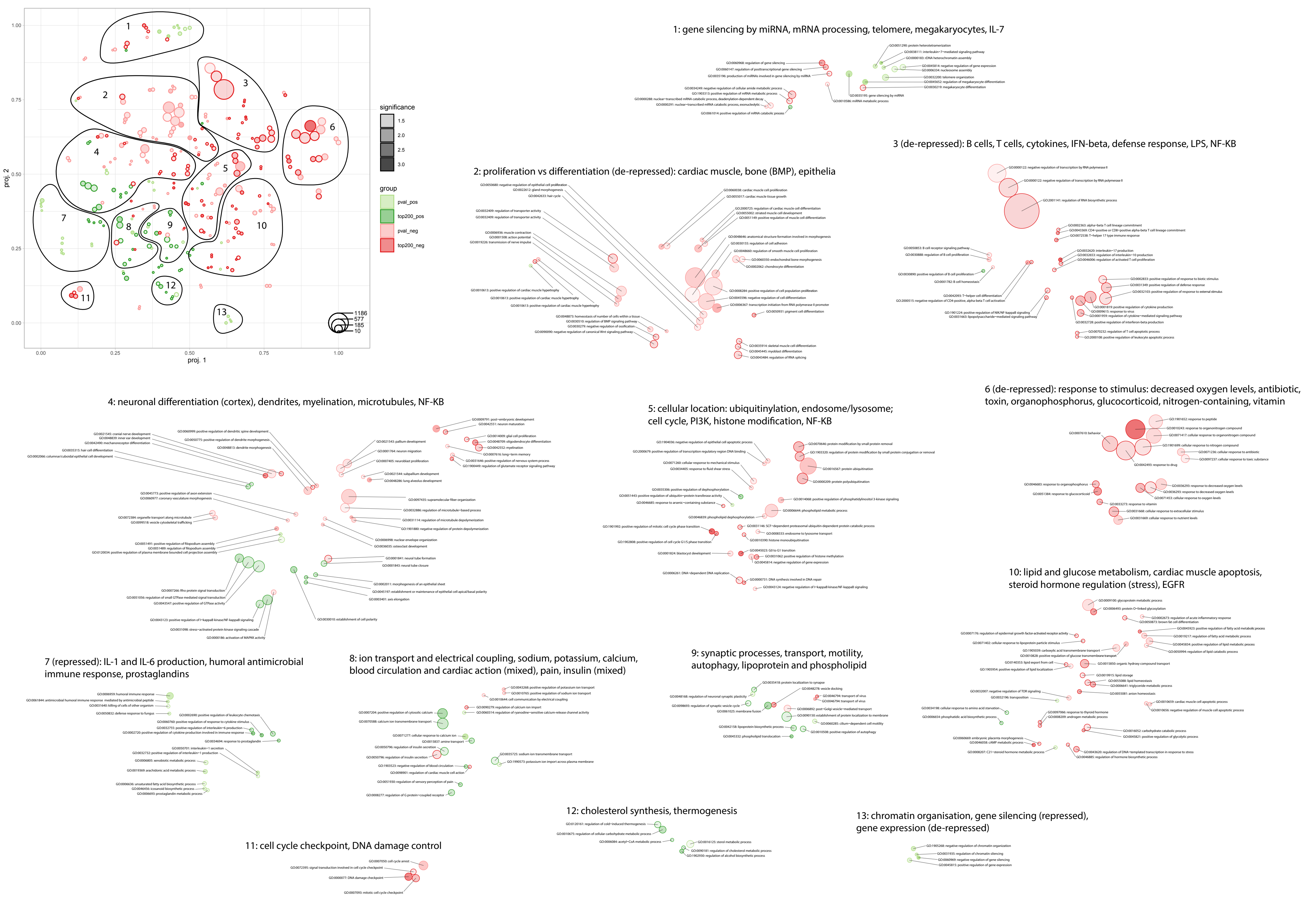

proj. 1
proj. 2
significance
1.5
2.0
2.5
3.0
group
pval_pos
top200_pos
pval_neg
top200_neg
1186
577
185
10
1: gene silencing by miRNA, mRNA processing, telomere, megakaryocytes, IL-7
2: proliferation vs differentiation (de-repressed): cardiac muscle, bone (BMP), epithelia
3 (de-repressed): B cells, T cells, cytokines, IFN-beta, defense response, LPS, NF-KB
4: neuronal differentiation (cortex), dendrites, myelination, microtubules, NF-KB
5: cellular location: ubiquitinylation, endosome/lysosome; cell cycle, PI3K, histone modification, NF-KB
6 (de-repressed): response to stimulus: decreased oxygen levels, antibiotic, toxin, organophosphorus, glucocorticoid, nitrogen-containing, vitamin
7 (repressed): IL-1 and IL-6 production, humoral antimicrobial immune response, prostaglandins
8: ion transport and electrical coupling, sodium, potassium, calcium, blood circulation and cardiac action (mixed), pain, insulin (mixed)
9: synaptic processes, transport, motility, autophagy, lipoprotein and phospholipid
10: lipid and glucose metabolism, cardiac muscle apoptosis, steroid hormone regulation (stress), EGFR
11: cell cycle checkpoint, DNA damage control
12: cholesterol synthesis, thermogenesis
13: chromatin organisation, gene silencing (repressed), gene expression (de-repressed)

**Figure S5 (on previous page)**

**Ontological enrichment and clustering of gene target sets of DE miRs show distinct perturbed pathways.** Gene target sets of positively and negatively perturbed miRs were subjected to gene ontology analysis separately, based on two scoring systems: “pval,” based on enrichment p-value in targeting permutation, and “top200”, based on the top 200 genes with highest amounts of miRs targeting each gene. Resulting GO terms were visualized using R/gsoap and clustered based on their shared genes (upper left). Circle sizes represent the number of genes in each term, their color depth denotes p-value (all $p < 0.05$). The single clusters were manually screened and annotated regarding their functions (right side).

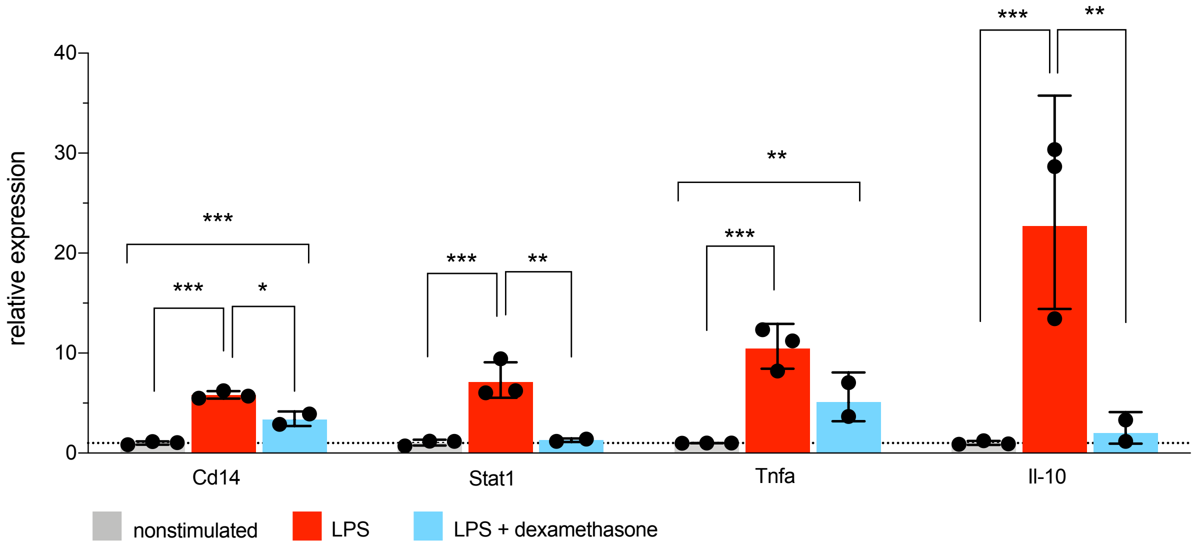


**Figure S6**

**Upregulated inflammatory signaling molecules and cytokines in LPS-stimulated murine RAW 264.7 cells.** A) Levels of cluster of differentiation 14 (Cd14), signal transducer and activator of transcription 1 (Stat1), tumor necrosis factor alpha (Tnfa) and interleukin 10 (Il-10) were measured with RT-qPCR using normalized expression (Ppia and 18SrRNA served as house-keeping), relative to the nonstimulated control group (line at mean normalized expression for the control group = 1), Each dot represents 2-4 technical replicates, ANOVA with Tukey post-hoc, * $p < 0.05$, ** $p < 0.01$, *** $p < 0.001$, bar graphs +/- SD(lg).

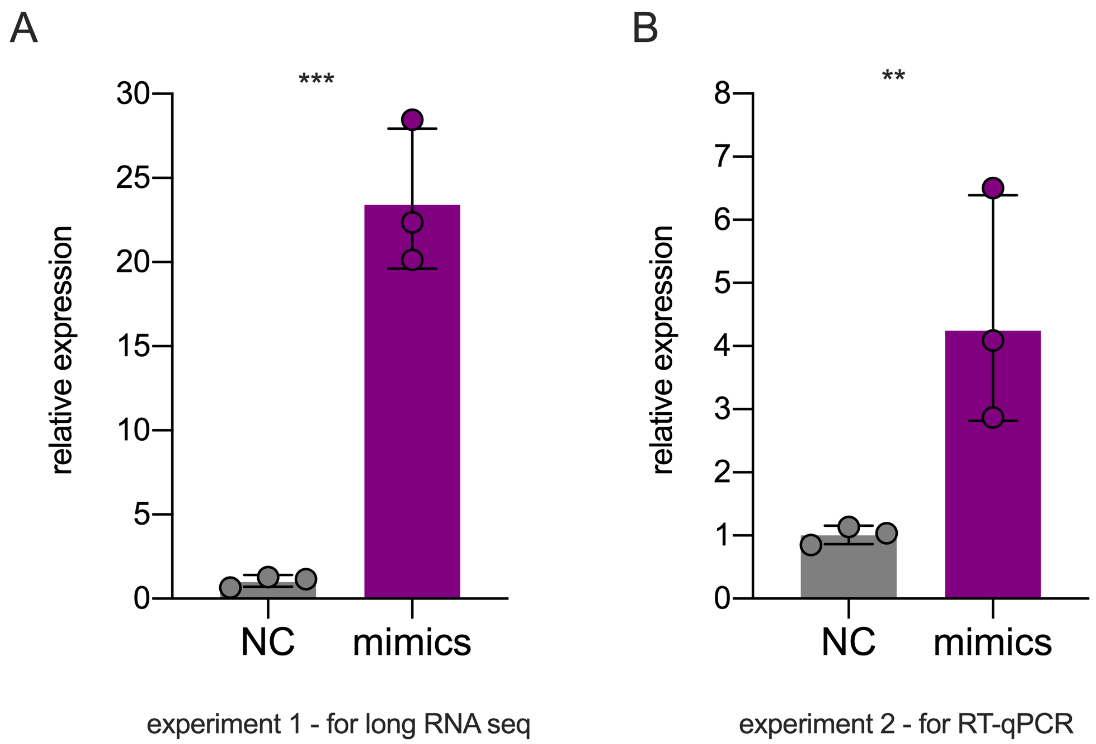


**Figure S7**

**tRF-22-WE8SPOX52 expression in the ssRNA mimics transfection experiments.** Shown is normalized expression of tRF-22-WE8SPOX52 and parental tRNA molecule (measurement without size selection) by RT-qPCR using Snord47 as house-keeping gene. A) Experiment 1 - samples subjected to long RNA sequencing. B) Experiment 2 - samples used for RT-qPCR measurements of Zbp1 expression. Each dot represents one technical replicate in the cell culture experiment. ** $p < 0.01$, *** $p < 0.001$, one-way ANOVA, bar graphs +/- SD(lg).

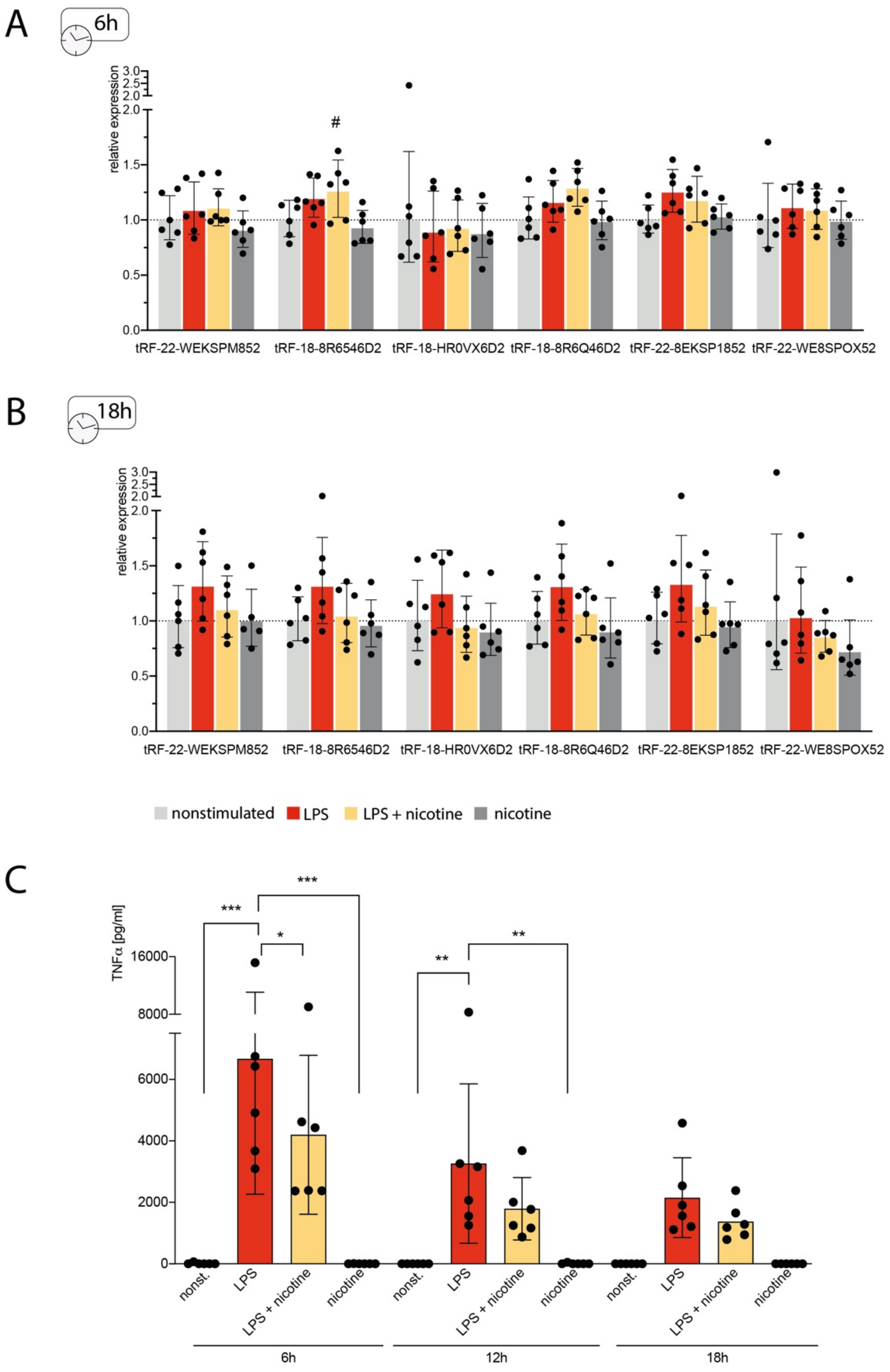


**Figure S8**

**LPS stimulation of human CD14+ cells yields transient nicotine-affected tRF changes**. The top 6 stroke-perturbed tRFs were quantified in human CD14+ cells following LPS stimulation with and without addition of nicotine; controls were nonstimulated cells and cells with nicotine alone (for setup, see Figure 6G in the main manuscript). A) Cells collected 6h after LPS stimulation showed significantly elevated tRF-18-8R6546D2 under LPS + nicotine exposure alone. B) At 18h after LPS stimulation, none of the top 6 stroke-perturbed tRFs showed significant changes, including LPS +nicotine challenged cells which showed upregulation at the 12h time point (see Figure 6H in the main manuscript). LPS-stimulated cells still showed the highest expression of tRFs, albeit not significant when compared to other groups. $^{\#}p < 0.05$ when compared to nicotine-treated cells. Each dot represents one donor, one-way ANOVA with Tukey post-hoc; bar graphs +/- SD(lg). C) LPS-stimulation led to significantly elevated TNF$\alpha$ levels in the supernatant of CD14+ cells (measured by ELISA). * $p<0.05$, ** $p<0.01$, *** $p<0.001$, two-way ANOVA on the influence of group and time variable on the TNF$\alpha$ levels. $F(3, 60) = 23.53$, $p <0.0001$ for the main effect “group” (nonstimulated, LPS, LPS + nicotine and nicotine), $F(2,60) = 7.56$ $p = 0.0012$ for the main effect “time”, $F(6, 60)$ $p=0.02$ for the interaction; with Dunnett’s post-hoc performing comparisons LPS group vs. others; bar graph +/- SD.

5’ AUCCCACCGCUGCCACCA 3’ trF-18-HR0VX6D2
5’ AUCCCACCACUGCCACCAU 3’ hsa-miR-1260b
5’ AUCCCACCUCUGCCACCA 3’ hsa-miR-1260a

**Figure S9**
**Sequence of tRF-18-HR0VX6D2 shows high similarities to two known miRs** (13)
hsa-miR-1260b differs from tRF-18-HR0VX6D2 by one nucleotide at position 9 and an additional nucleotide at the 3'-end, has-miR-1260a differs from tRF-18-HR0VX6D2 only at position 9.

| GO.ID | Term | Annotated | Significant | Expected | Rank in classic | classic | weight |
|---|---|---|---|---|---|---|---|
| GO:0001819 | positive regulation of cytokine producti... | 66 | 14 | 4.38 | 42 | 6E-05 | 0.00017 |
| GO:0032479 | regulation of type I interferon producti... | 29 | 8 | 1.93 | 52 | 0.00039 | 0.00039 |
| GO:0001818 | negative regulation of cytokine producti... | 51 | 11 | 3.39 | 51 | 0.00034 | 0.00057 |
| GO:0009617 | response to bacterium | 95 | 18 | 6.31 | 35 | 2.5E-05 | 0.00059 |
| GO:0045087 | innate immune response | 151 | 38 | 10.03 | 1 | 9.1E-15 | 0.00197 |
| GO:0098586 | cellular response to virus | 15 | 5 | 1 | 65 | 0.00208 | 0.00208 |
| GO:0046683 | response to organophosphorus | 22 | 6 | 1.46 | 68 | 0.00233 | 0.00233 |
| GO:0002753 | cytoplasmic pattern recognition receptor... | 10 | 4 | 0.66 | 70 | 0.00284 | 0.00284 |
| GO:0048661 | positive regulation of smooth muscle cel... | 16 | 5 | 1.06 | 72 | 0.00286 | 0.00286 |
| GO:0060760 | positive regulation of response to cytok... | 16 | 5 | 1.06 | 73 | 0.00286 | 0.00286 |
| GO:0014074 | response to purine-containing compound | 23 | 6 | 1.53 | 75 | 0.00299 | 0.00299 |
| GO:0032649 | regulation of interferon-gamma productio... | 17 | 5 | 1.13 | 80 | 0.00384 | 0.00384 |
| GO:0016525 | negative regulation of angiogenesis | 12 | 4 | 0.8 | 88 | 0.00603 | 0.00603 |
| GO:0034446 | substrate adhesion-dependent cell spread... | 13 | 4 | 0.86 | 93 | 0.00827 | 0.00827 |
| GO:0032496 | response to lipopolysaccharide | 46 | 8 | 3.06 | 96 | 0.00917 | 0.00917 |
| GO:0009063 | cellular amino acid catabolic process | 14 | 4 | 0.93 | 100 | 0.01099 | 0.01099 |
| GO:0031349 | positive regulation of defense response | 58 | 11 | 3.85 | 61 | 0.00109 | 0.01236 |
| GO:0002576 | platelet degranulation | 22 | 5 | 1.46 | 102 | 0.01252 | 0.01252 |
| GO:0010469 | regulation of signaling receptor activit... | 49 | 8 | 3.26 | 103 | 0.01340 | 0.01340 |
| GO:0070887 | cellular response to chemical stimulus | 404 | 54 | 26.84 | 19 | 6.2E-09 | 0.01563 |
| GO:0030168 | platelet activation | 32 | 6 | 2.13 | 111 | 0.01641 | 0.01641 |
| GO:0034109 | homotypic cell-cell adhesion | 16 | 4 | 1.06 | 112 | 0.01802 | 0.01802 |
| GO:0050731 | positive regulation of peptidyl-tyrosine... | 25 | 5 | 1.66 | 117 | 0.02152 | 0.02152 |
| GO:0048469 | cell maturation | 17 | 4 | 1.13 | 118 | 0.02238 | 0.02238 |
| GO:0097696 | STAT cascade | 17 | 4 | 1.13 | 119 | 0.02238 | 0.02356 |
| GO:0043330 | response to exogenous dsRNA | 10 | 3 | 0.66 | 123 | 0.02429 | 0.02429 |
| GO:0071695 | anatomical structure maturation | 18 | 4 | 1.2 | 127 | 0.02734 | 0.02734 |
| GO:0050680 | negative regulation of epithelial cell p... | 19 | 4 | 1.26 | 139 | 0.03290 | 0.03290 |
| GO:0007267 | cell-cell signaling | 146 | 11 | 9.7 | 649 | 0.37575 | 0.03503 |
| GO:0001936 | regulation of endothelial cell prolifera... | 20 | 4 | 1.33 | 145 | 0.03907 | 0.03907 |
| GO:0006906 | vesicle fusion | 20 | 4 | 1.33 | 146 | 0.03907 | 0.03907 |
| GO:0007160 | cell-matrix adhesion | 20 | 4 | 1.33 | 147 | 0.03907 | 0.03907 |
| GO:0030856 | regulation of epithelial cell differenti... | 12 | 3 | 0.8 | 151 | 0.04041 | 0.04041 |
| GO:0034113 | heterotypic cell-cell adhesion | 12 | 3 | 0.8 | 152 | 0.04041 | 0.04041 |
| GO:0042130 | negative regulation of T cell proliferat... | 12 | 3 | 0.8 | 153 | 0.04041 | 0.04041 |
| GO:0051668 | localization within membrane | 12 | 3 | 0.8 | 154 | 0.04041 | 0.04041 |

**Table S1**
**Post-stroke DE genes are enriched in circulation- and immunity- relelated pathways.**
Full list of GO terms used for generation of tSNE in Figure 4 (main manuscript).
